# Anti-SARS-CoV-2 antibody containing plasma improves outcome in patients with hematologic or solid cancer and severe COVID-19 via increased neutralizing antibody activity – a randomized clinical trial

**DOI:** 10.1101/2022.10.10.22280850

**Authors:** C.M. Denkinger, M. Janssen, U. Schäkel, J. Gall, A. Leo, P. Stelmach, S. F. Weber, J. Krisam, L. Baumann, J. Stermann, U. Merle, M. A. Weigand, C. Nusshag, L. Bullinger, J.F. Schrezenmeier, M. Bornhäuser, N. Alakel, O. Witzke, T. Wolf, M. J.G.T. Vehreschild, S. Schmiedel, M. M. Addo, F. Herth, M. Kreuter, P.-R. Tepasse, B. Hertenstein, M. Hänel, A. Morgner, M. Kiehl, O. Hopfer, M.-A. Wattad, C. C. Schimanski, C. Celik, T. Pohle, M. Ruhe, W. V. Kern, A. Schmitt, H.M. Lorenz, M. Souto-Carneiro, M. Gaeddert, N. Halama, S. Meuer, H.G. Kräusslich, B. Müller, P. Schnitzler, S. Parthé, R. Bartenschlager, M. Gronkowski, J. Klemmer, M. Schmitt, P. Dreger, K. Kriegsmann, R. F. Schlenk, C. Müller-Tidow

**Affiliations:** Division of Infectious Disease and Tropical Medicine, Department of Infectious Diseases, Heidelberg University Hospital, Heidelberg, Germany; Department of Internal Medicine V, Heidelberg University Hospital, Heidelberg, Germany; NCT-Trial Center, National Center of Tumor Diseases, Heidelberg University Hospital and German Cancer Research Center, Heidelberg, Germany; Institute for Clinical Transfusion Medicine and Cell Therapy Heidelberg, Heidelberg, Germany; Institute of Medical Biometry (IMBI), University of Heidelberg, Heidelberg, Germany; Department of Internal Medicine IV, Heidelberg University Hospital, Heidelberg, Heidelberg, Germany; Department of Anesthesiology, Heidelberg University Hospital, Heidelberg, Germany; Department of Nephrology, University of Heidelberg, Heidelberg; Department of Hematology, Oncology and Tumor Immunology, Charité-Universita□tsmedizin Berlin, corporate member of Freie Universita□t Berlin, Humboldt-Universita□t zu Berlin, Berlin, Germany; Department of Internal Medicine I, University Hospital and Faculty of Medicine Carl Gustav Carus of TU Dresden, Dresden, Germany; Department of Infectious Diseases, West German Centre of Infectious Diseases, University Hospital Essen, University Duisburg-Essen, Essen, Germany; Department of Internal Medicine, Infectious Diseases, University Hospital Frankfurt, Goethe University Frankfurt, Frankfurt am Main, Germany; I Department of Medicine, University Medical Center Hamburg-Eppendorf, Hamburg, Germany; Pneumology and Critical Care Medicine Thoraxklinik, University of Heidelberg and Translational Lung Research Center (TIRC) Heidelberg, Germany; Center for Interstitial and Rare Lung Diseases, Pneumology and Critical Care Medicine, Thoraxklinik, University of Heidelberg and German Center for Lung Research (DZL), Heidelberg, Germany; Department of Medicine B, Gastroenterology and Hepatology, University Hospital Münster, Münster, Germany; Medical Department I, Klinikum Bremen-Mitte, Bremen, Germany; Department of Internal Medicine III, Klinikum Chemnitz gGmbH, Chemnitz, Germany; Department I of Internal Medicine, Frankfurt (Oder) General Hospital, Frankfurt (Oder), Germany; Department of Hematology, Oncology, Palliative Care and Stem Cell Transplantation, Klinikum Hochsauerland GmbH, Meschede, Germany; Department of Internal Medicine II, Klinikum Darmstadt GmbH, Darmstadt, Germany; Department of Internal Medicine I, Klinikum Herford, Herford, Germany; Department of Medicine II, Division of Infectious Diseases and Travel Medicine, University Medical Centre Freiburg, Freiburg, Germany; Department of Medical Oncology, National Center for Tumor Diseases, Heidelberg University Hospital, Heidelberg, Germany; Department of Infectious Diseases, Virology, Heidelberg University Hospital, Heidelberg, Germany; Department of Infectious Diseases, Molecular Virology, Heidelberg University Hospital, Heidelberg, Germany; National Center for Tumor Diseases (NCT) Heidelberg, Heidelberg, Germany; German Center for Infection Research (DZIF), partner site Heidelberg University Hospital, Heidelberg; partner site Hamburg-Lübeck-Borstel-Riems, Germany; Department of Translational Immunotherapy (D240), German Cancer Research Center (DKFZ), Heidelberg, Germany; Helmholtz Institute for Translational Oncology (HI-TRON), Mainz, Germany; University Medical Center Hamburg-Eppendorf, Institute for Infection Research and Vaccine Development (IIRVD); Hamburg, Germany

**Keywords:** SARS-CoV-2, plasma therapy, cancer, immunodeficiency, randomized-controlled trial

## Abstract

Cancer patients are at high risk of severe COVID-19 with high morbidity and mortality. Further, impaired humoral response renders SARS-CoV-2 vaccines less effective and treatment options are scarce. Randomized trials using convalescent plasma are missing for high-risk patients. Here, we performed a multicenter trial (https://www.clinicaltrialsregister.eu/ctr-search/trial/2020-001632-10/DE) in hospitalized patients with severe COVID-19 within four risk groups (1, cancer; 2, immunosuppression; 3, lab-based risk factors; 4, advanced age) randomized to standard of care (CONTROL) or standard of care plus convalescent/vaccinated anti-SARS-CoV-2 plasma (PLASMA). For the four groups combined, PLASMA did not improve clinically compared to CONTROL (HR 1.29; *p*=0.205). However, cancer patients experienced shortened median time to improvement (HR 2.50, *p*=0.003) and superior survival in PLASMA vs. CONTROL (HR 0.28; *p*=0.042). Neutralizing antibody activity increased in PLASMA but not in CONTROL cancer patients (*p*=0.001). Taken together, convalescent/vaccinated plasma may improve COVID-19 outcome in cancer patients unable to intrinsically generate an adequate immune response.

## Introduction

COVID-19 associated risk of death is particularly high for patients with hematologic or solid cancer^1-3^, advanced age^4,5^ and other conditions^6,7^. Both humoral^8^ and cellular^9^ immunodeficiency contribute to unfavorable outcomes. Despite, SARS-CoV-2 vaccine availability and waning vaccine efficacy in these patients remain concerning^10,11^.

Few therapies improve outcomes in severe COVID-19 with impaired oxygenation^12^. Monoclonal antibodies as pre- or post-exposure prophylaxis or as early treatment can reduce the risk of severe COVID-19^13,14^. Evidence for the benefit of monoclonal antibodies in patients requiring oxygen supplementation is missing^15^ or pending^16^.

Clinical trials on convalescent plasma therapy for COVID-19 were mostly negative^17-24^. Relevant determinants causing heterogeneity in plasma efficacy were: (1) Timing from disease onset to therapy initiation, with early therapy being most effective^18^ and (2) Titers of neutralizing antibodies^18,23,24^. Still, it is unknown whether patients without sufficient antibody response benefit from therapy with plasma from convalescent or vaccinated donors, but several subgroup analyses have pointed towards better outcomes with plasma therapy. In a Bayesian re-analysis of the RECOVERY trial, the subgroup of patients who had not yet developed an antibody response to SARS-CoV-2 appeared to have slightly better outcomes when treated with convalescent plasma^25^. A similar subgroup analysis of the REMAP-CAP trial pointed towards a potential benefit for immunosuppressed patients^26^. Two observational propensity-score matched cohort studies showed a marked reduction in mortality despite in parts later disease stages at time of transfusion^27,28^.

Here, we performed a randomized controlled clinical trial with convalescent/vaccinated plasma in high-risk patients including cancer patients with severe COVID-19 and analyzed the association between plasma therapy and response of neutralizing antibody titers in plasma recipients.

## Results

### Trial Population

A total of 136 patients meeting eligibility criteria were randomized (Figure 1). Inclusion criteria were: (1) polymerase chain reaction (PCR) confirmed infection with SARS-CoV-2 in a respiratory tract sample; (2) oxygen saturation (SaO2) on ambient air of ≤94% or a partial oxygen pressure (PaO2) – inspired oxygen fraction (FiO2) ratio of <300mmHg; (3) provision of written informed consent; and (4) meeting at least one high-risk criterion to define the patient group (supplement: study protocol):

**Figure 1:**
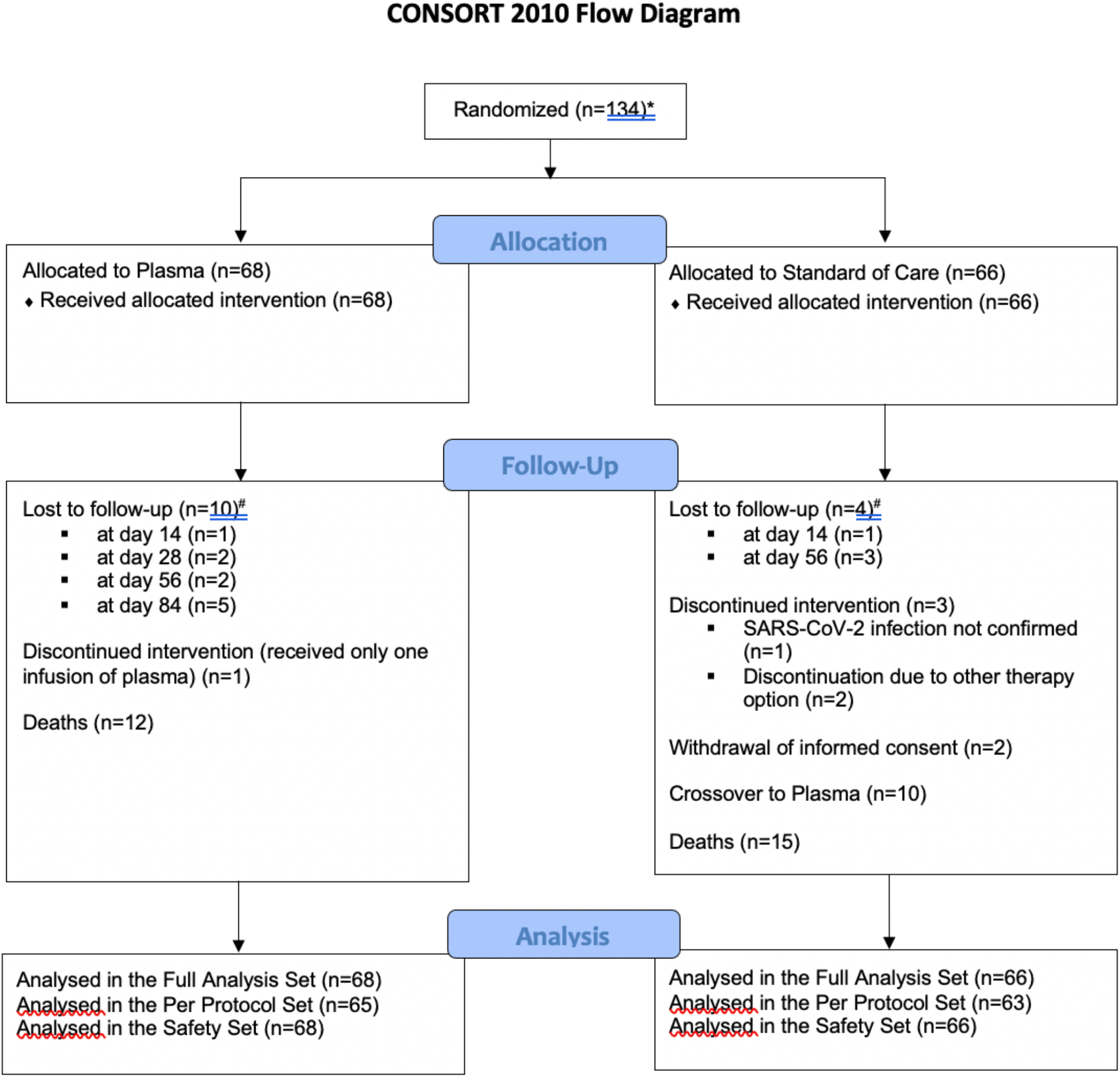
Consort Diagram. Patient flow within the RECOVER trial.

⍰ Group-1 – ‘cancer’: patients with pre-existing or concurrent hematological cancer and/or active cancer therapy for any cancer (incl. chemo-, radio- and surgical treatments) within the past 24 months.
⍰ Group-2 – ‘immunosuppression’: patients under chronic immunosuppression either pharmacological and/or due to underlying diseases not meeting Group-1 criteria.
⍰ Group-3 – ‘lymphopenia/elevated D-dimers’: patients aged >50 and ≤75 years not meeting Group-1 or -2 criteria who had lymphopenia (<0.8 G/l) and/or D-dimers (>1µg/ml).
⍰ Group-4 – ‘age >75 years’: patients aged >75 years not meeting Group-1, -2 or -3 criteria.

Two patients were excluded due to absence of a signed informed consent and withdrawal of consent after signature, respectively. Thus, 134 patients were enrolled and randomized between September 3, 2020 and January 20, 2022. 68 patients were assigned to PLASMA and 66 to CONTROL treatment. Eligible patients underwent randomization into the experimental (PLASMA) or control (CONTROL) arm at a 1:1 ratio using block randomization for the patient group strata defined above (groups-1 to -4). Patients in PLASMA received at least one unit of AB0-compatible plasma, and 10 CONTROL patients crossed over to PLASMA at day 10 after randomization. Plasma donor eligibility required high titers of neutralizing antibody activity in a live virus neutralization assay (titers ≥1:80; less than 20% of potential donors) (Supplementary text S2, S3, Figure S6, Table S4). Recruitment was stopped on January 20th, 2022, after enrolment of 77% of the target population (Methods). Average age was 69 years (range, 36-95 years) (Table 1; Table S5) and 43 patients were female (32.1%). ECOG performance status (median 2), clinical frailty scale (median 3) and time from symptom onset to randomization (median 7.0) were similar in both arms. The allocation of patients to the predefined high-risk patient groups were: group-1 42% (*n*=56, Figure S1), group-2 12% (*n*=16), group-3 27% (*n*=36), group-4 19% (*n*=26) (Table 1; group details in Table S5). The most common cancers were B-cell malignancies (*n*=20), acute myeloid leukemia/ myelodysplastic syndrome (*n*=12) and myeloma (*n*=11), and solid cancer (*n*=9). Two patients suffered from Hodgkin’s lymphoma and one patient each from chronic myeloid leukemia or T-cell lymphoma (Table 1). The most common cause for chronic immunosuppression in group-2 was solid organ transplantation (*n*=12). In group-3, 27 patients showed lymphopenia and 21 patients elevated d-dimers, both criteria were present in 12 patients.

**Table 1:**
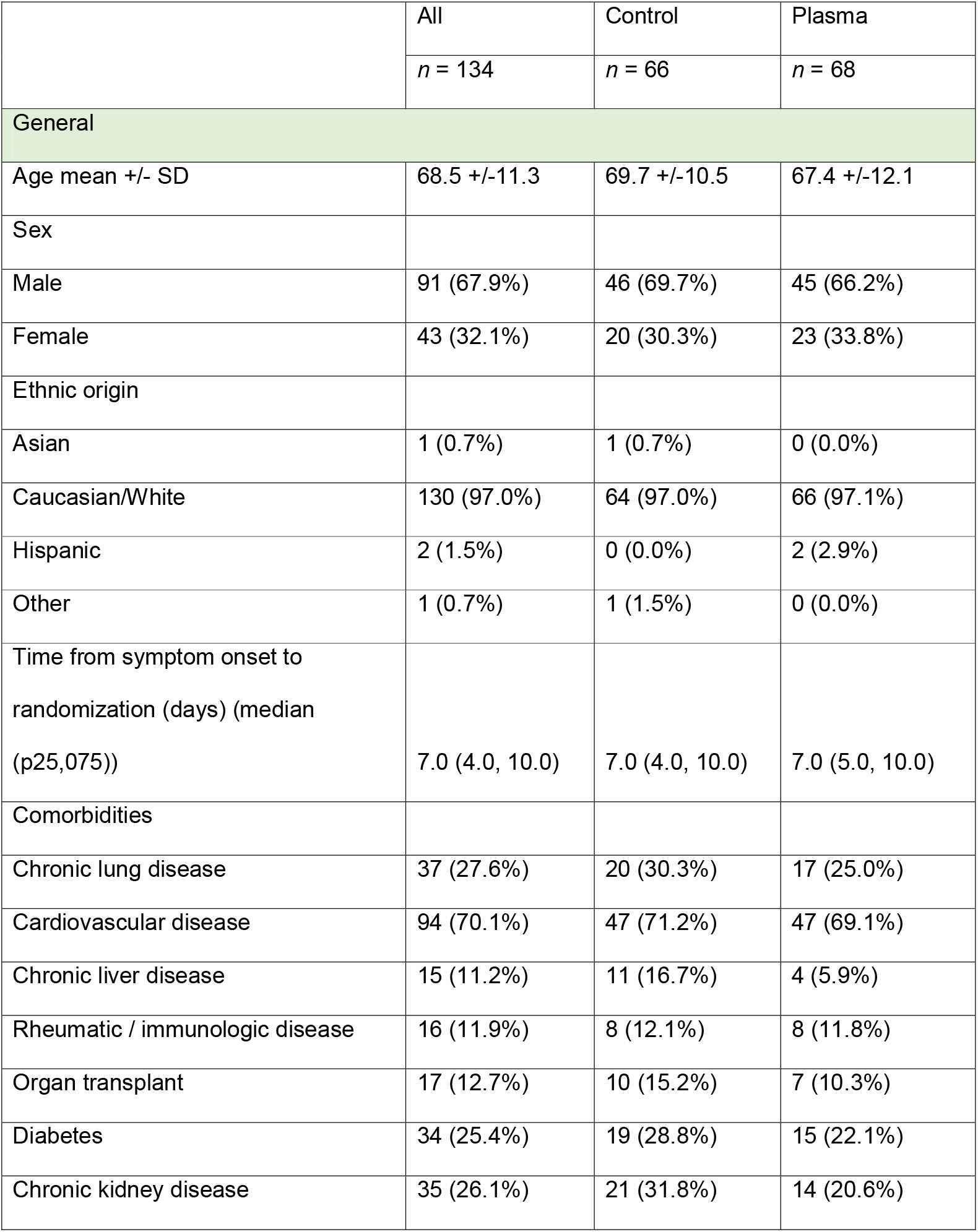
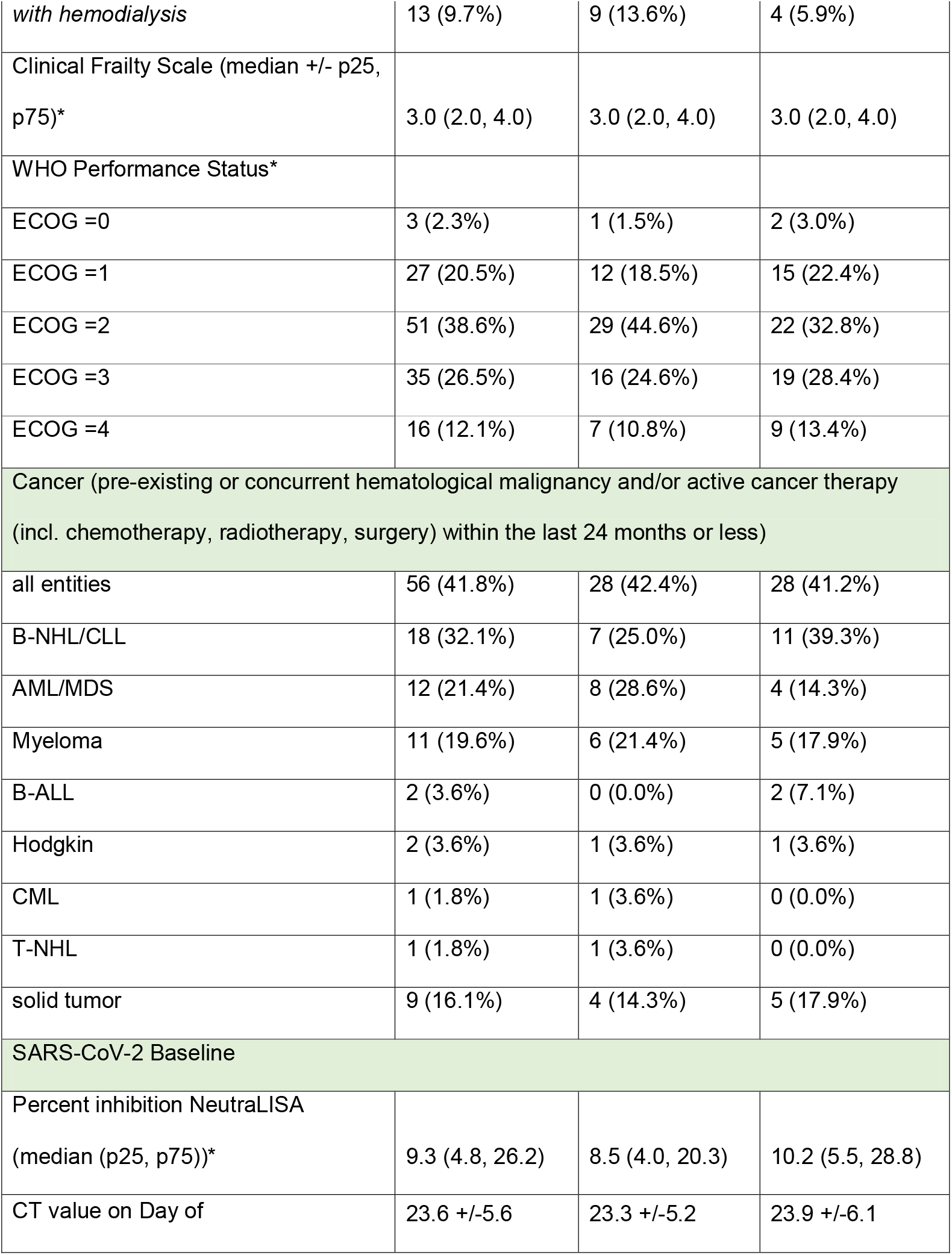
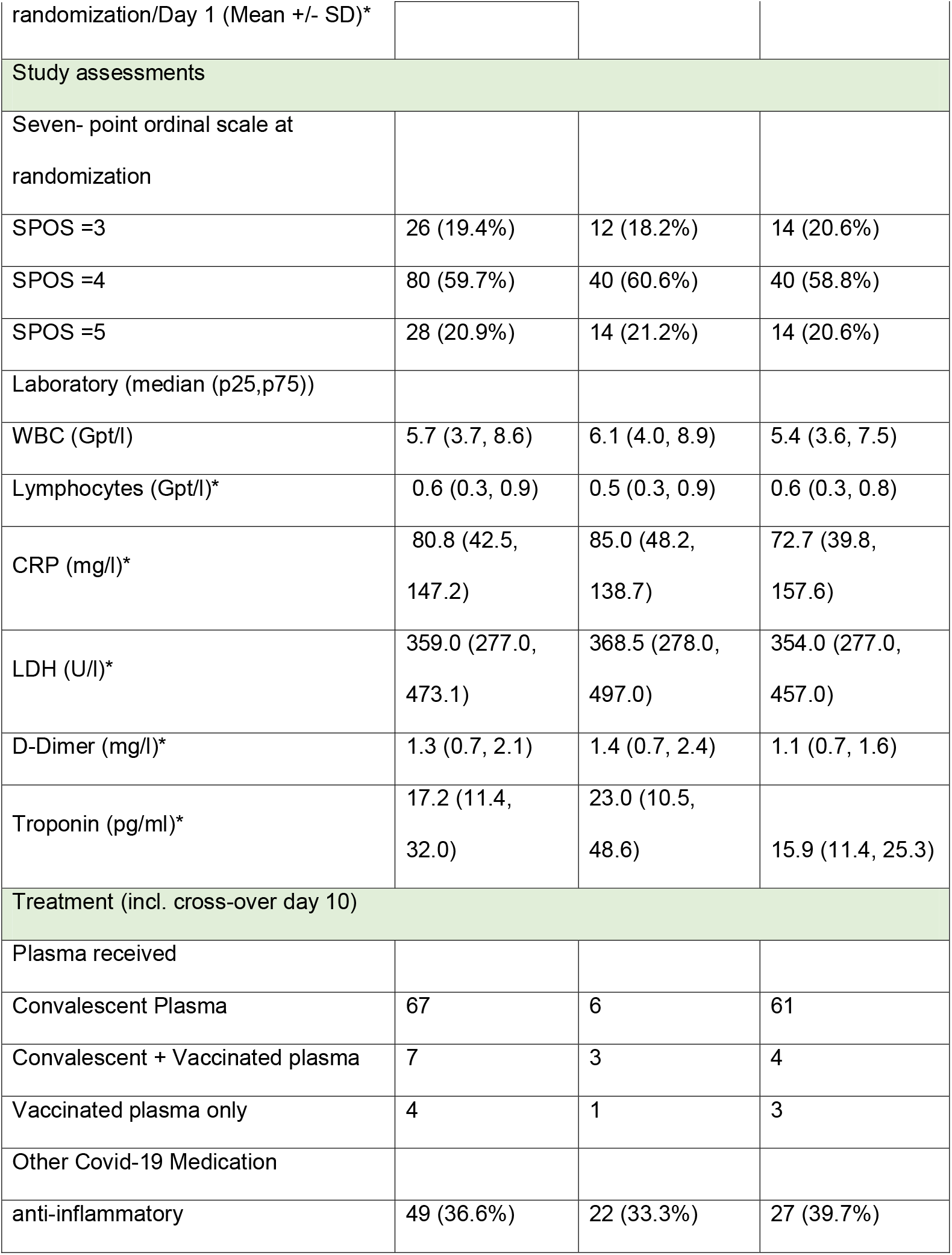
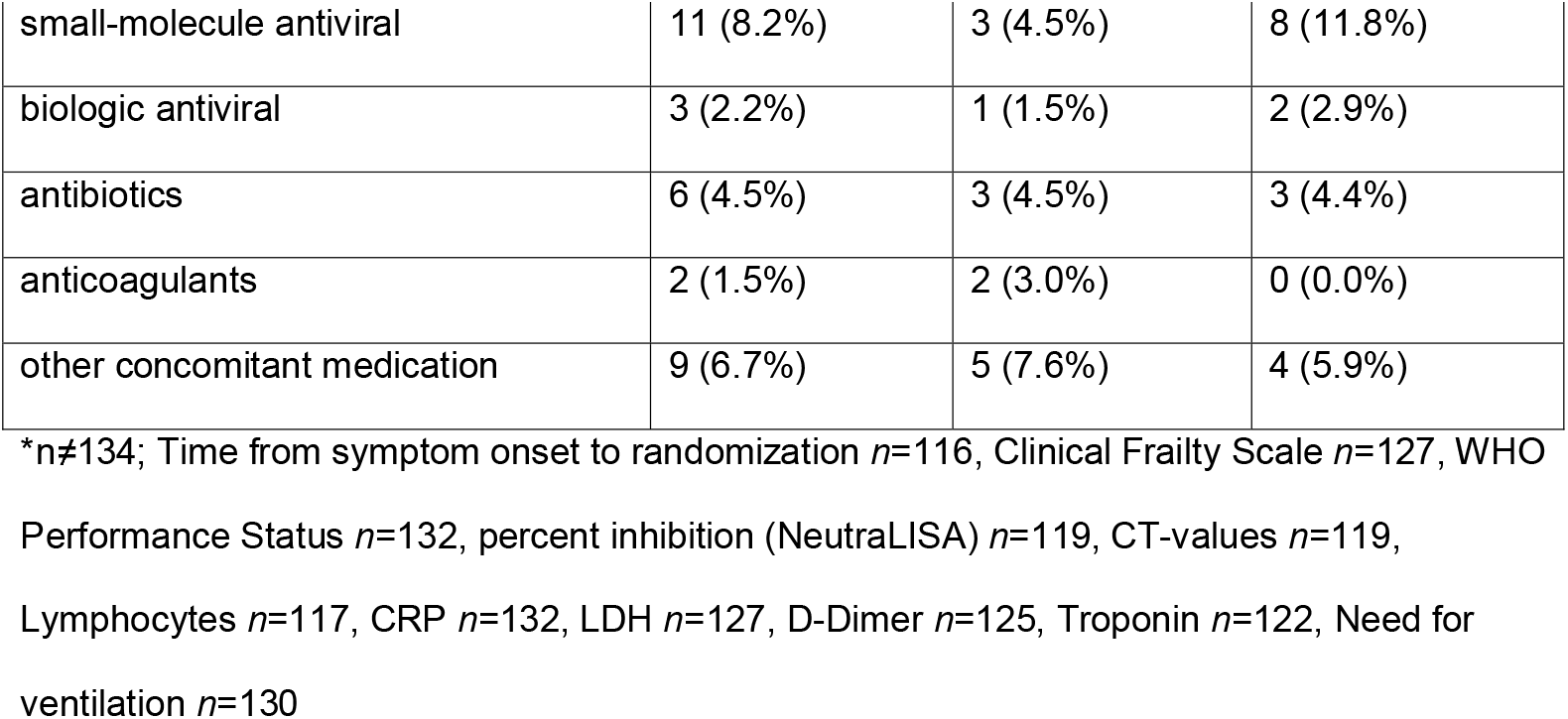
Patient and treatment characteristics.

### Follow-up and primary endpoint

A clinical seven point ordinal scale (7POS)^29,30^ was determined daily, which was defined as: 1, not hospitalized with resumption of normal activities; 2, not hospitalized, but unable to resume normal activities; 3, hospitalized, not requiring supplemental oxygen; 4, hospitalized, requiring supplemental oxygen; 5, hospitalized, requiring nasal high-flow oxygen therapy, noninvasive mechanical ventilation, or both; 6, hospitalized, requiring ECMO, invasive mechanical ventilation, or both; and 7, death^31^. At baseline, the 7POS was in median 4 (range 3-5) and oxygen supplementation (through nasal cannula or high-flow oxygen therapy) was required in *n*=108 (80.6%) patients with equal distribution in both arms.

In the full analysis set, median time from randomization to improvement of 2 points on the 7-point ordinal scale or live hospital discharge in PLASMA and CONTROL was 12.5 days (95%-confidence interval (CI), 10-17) in PLASMA and 18 days (95%-CI, 11-28) in CONTROL (hazard ratio (HR) 1.29; 95%-CI, 0.86-1.93, log-rank *p*=0.205), (Figure 2A&B). Pre-specified subgroup analyses revealed benefit in patients with cancer (group-1, *n*=56). For cancer patients, median time to improvement was 13 days (95%-CI, 7-14) for PLASMA and 31 days (95%-CI, 15-NA) for CONTROL, (HR 2.50; 95%-CI, 1.34-4.79, log-rank *p*=0.003, Figure 2B&C). Given potential confounders in age and gender distributions between the PLASMA and CONTROL arm, we adjusted for these variables in a sensitivity analysis. This resulted in a similar HR in group-1 (HR 2.79; 95%-CI, 1.35-5.94) supporting the beneficial role of plasma for cancer patients. No significant differences between arms were observed in groups-2 to -4 (Figure 2B&D, Figure S2, Table 2).

**Table 2:**
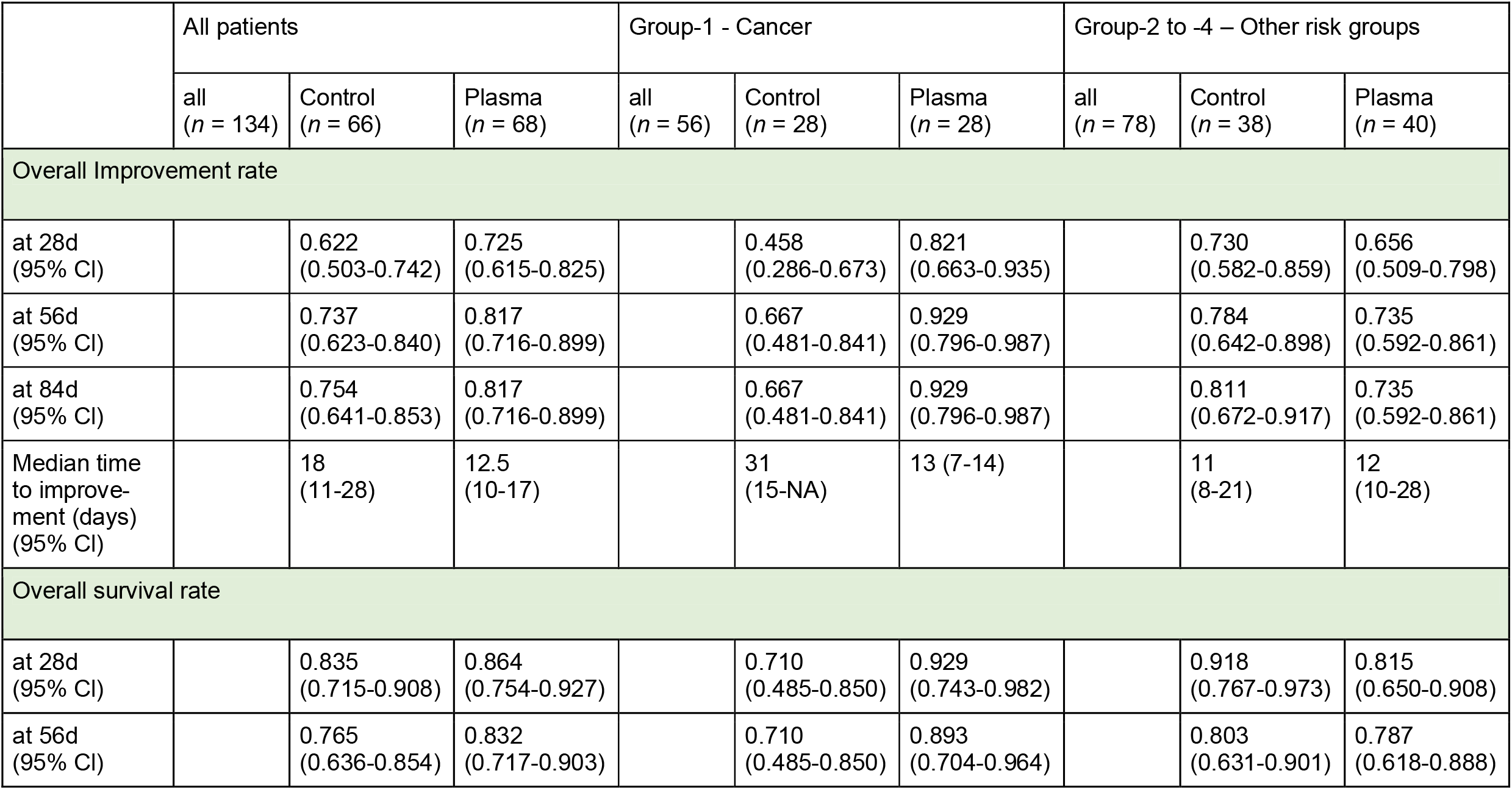
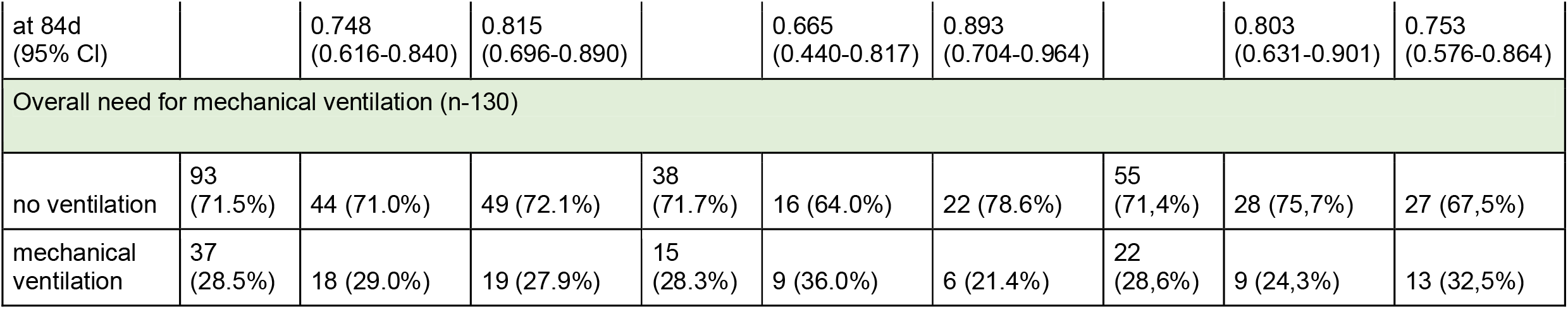
Outcome data overall and in group-1 and group-2 to -4

|  | All patients |  |  | Group-1 - Cancer |  |  | Group-2 to -4 – Other risk groups |  |  |
| --- | --- | --- | --- | --- | --- | --- | --- | --- | --- |
|  | all<br>(n = 134) | Control<br>(n = 66) | Plasma<br>(n = 68) | all<br>(n = 56) | Control<br>(n = 28) | Plasma<br>(n = 28) | all<br>(n = 78) | Control<br>(n = 38) | Plasma<br>(n = 40) |
| Overall Improvement rate |  |  |  |  |  |  |  |  |  |
| at 28d<br>(95% CI) |  | 0.622<br>(0.503-0.742) | 0.725<br>(0.615-0.825) |  | 0.458<br>(0.286-0.673) | 0.821<br>(0.663-0.935) |  | 0.730<br>(0.582-0.859) | 0.656<br>(0.509-0.798) |
| at 56d<br>(95% CI) |  | 0.737<br>(0.623-0.840) | 0.817<br>(0.716-0.899) |  | 0.667<br>(0.481-0.841) | 0.929<br>(0.796-0.987) |  | 0.784<br>(0.642-0.898) | 0.735<br>(0.592-0.861) |
| at 84d<br>(95% CI) |  | 0.754<br>(0.641-0.853) | 0.817<br>(0.716-0.899) |  | 0.667<br>(0.481-0.841) | 0.929<br>(0.796-0.987) |  | 0.811<br>(0.672-0.917) | 0.735<br>(0.592-0.861) |
| Median time<br>to improve-<br>ment (days)<br>(95% CI) |  | 18<br>(11-28) | 12.5<br>(10-17) |  | 31<br>(15-NA) | 13 (7-14) |  | 11<br>(8-21) | 12<br>(10-28) |
| Overall survival rate |  |  |  |  |  |  |  |  |  |
| at 28d<br>(95% CI) |  | 0.835<br>(0.715-0.908) | 0.864<br>(0.754-0.927) |  | 0.710<br>(0.485-0.850) | 0.929<br>(0.743-0.982) |  | 0.918<br>(0.767-0.973) | 0.815<br>(0.650-0.908) |
| at 56d<br>(95% CI) |  | 0.765<br>(0.636-0.854) | 0.832<br>(0.717-0.903) |  | 0.710<br>(0.485-0.850) | 0.893<br>(0.704-0.964) |  | 0.803<br>(0.631-0.901) | 0.787<br>(0.618-0.888) |
| at 84d<br>(95% CI) |  | 0.748<br>(0.616-0.840) | 0.815<br>(0.696-0.890) |  | 0.665<br>(0.440-0.817) | 0.893<br>(0.704-0.964) |  | 0.803<br>(0.631-0.901) | 0.753<br>(0.576-0.864) |
| Overall need for mechanical ventilation (n=130) |  |  |  |  |  |  |  |  |  |
| no ventilation | 93<br>(71.5%) | 44 (71.0%) | 49 (72.1%) | 38<br>(71.7%) | 16 (64.0%) | 22 (78.6%) | 55<br>(71,4%) | 28 (75,7%) | 27 (67,5%) |
| mechanical<br>ventilation | 37<br>(28.5%) | 18 (29.0%) | 19 (27.9%) | 15<br>(28.3%) | 9 (36.0%) | 6 (21.4%) | 22<br>(28,6%) | 9 (24,3%) | 13 (32,5%) |

**Figure 2:**
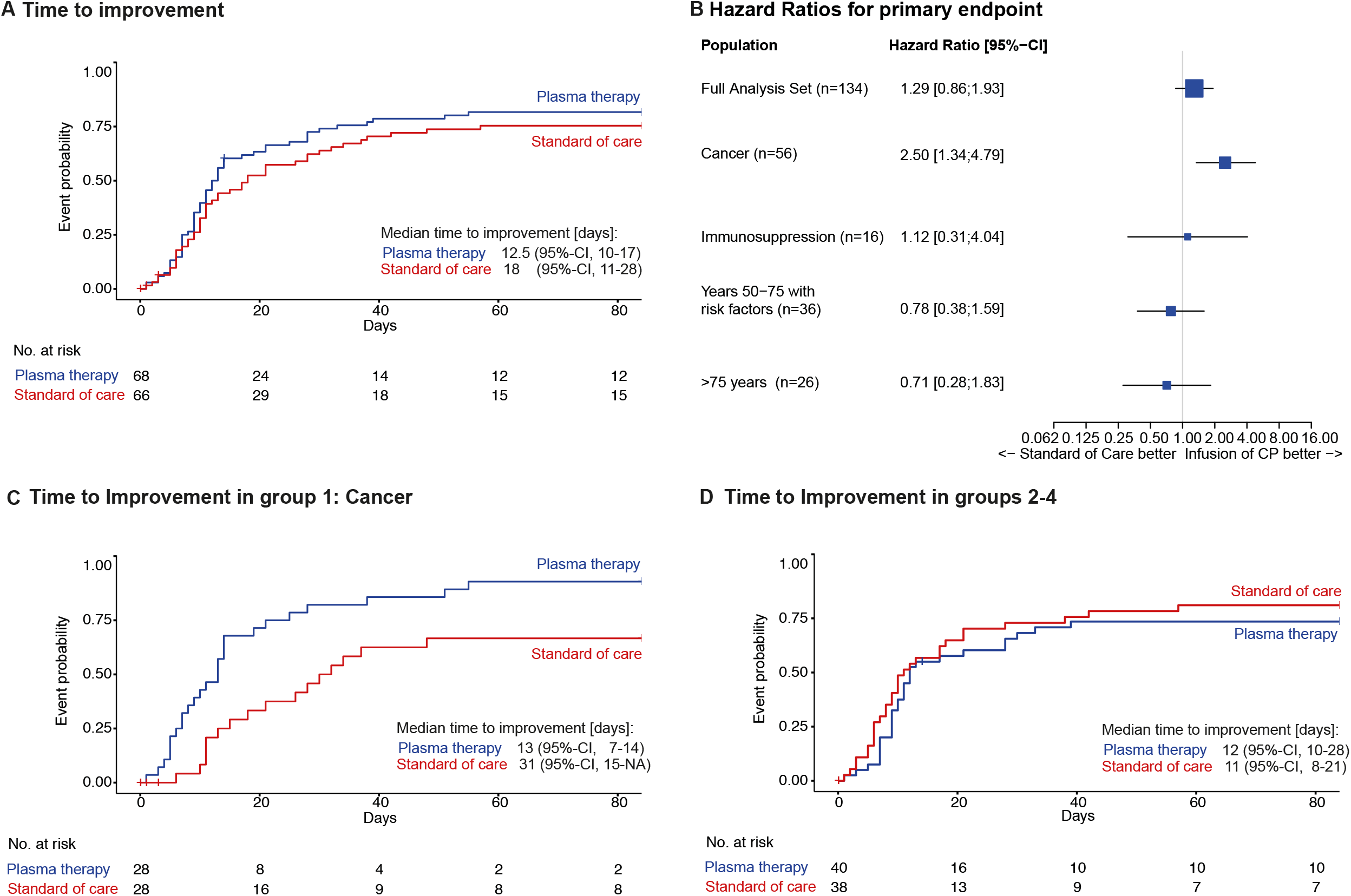
Primary endpoint - Time to discharge or improvement of 2 points in the 7-point ordinal scale or live hospital discharge. A: Kaplan Meier curve for primary endpoint of 2-point improvement on 7-point ordinal scale or live hospital discharge for overall study cohort (group-1 to 4) by PLASMA (blue) and CONTROL (red) with number of subjects at risk; log-rank *p*=0.205. B: Forest plot with hazard ratios (HR) for primary endpoint overall (full analysis set) and by predefined subgroups, 95% confidence intervals in brackets C: Kaplan Meier curve for primary endpoint for group-1 by PLASMA (blue) and CONTROL (red) with number of subjects at risk; log-rank *p*=0.003. D: Kaplan Meier curve for primary endpoint for combined groups-2 to -4 by PLASMA (blue) and CONTROL (red) with number of subjects at risk. See Supplementary Figure S3 for separate data of groups 2-4; log-rank *p*=0.3902.

### Overall Survival and other Secondary Endpoints

Overall, n=27 patients died, and no significant difference was seen for overall survival according to randomization (HR 0.72; 95%-CI, 0.33-1.55; log rank *p*=0.403) (Figure 2A). In the cancer group (group-1), improved overall survival was observed in PLASMA compared to CONTROL (HR 0.28; 95%-CI, 0.06-0.96; log-rank *p*=0.042) (Figure 3B&C). Treatment arms of groups 2-4 did not differ in survival (Figure 2B&D and Figure S3, Table 2).

**Figure 3.**
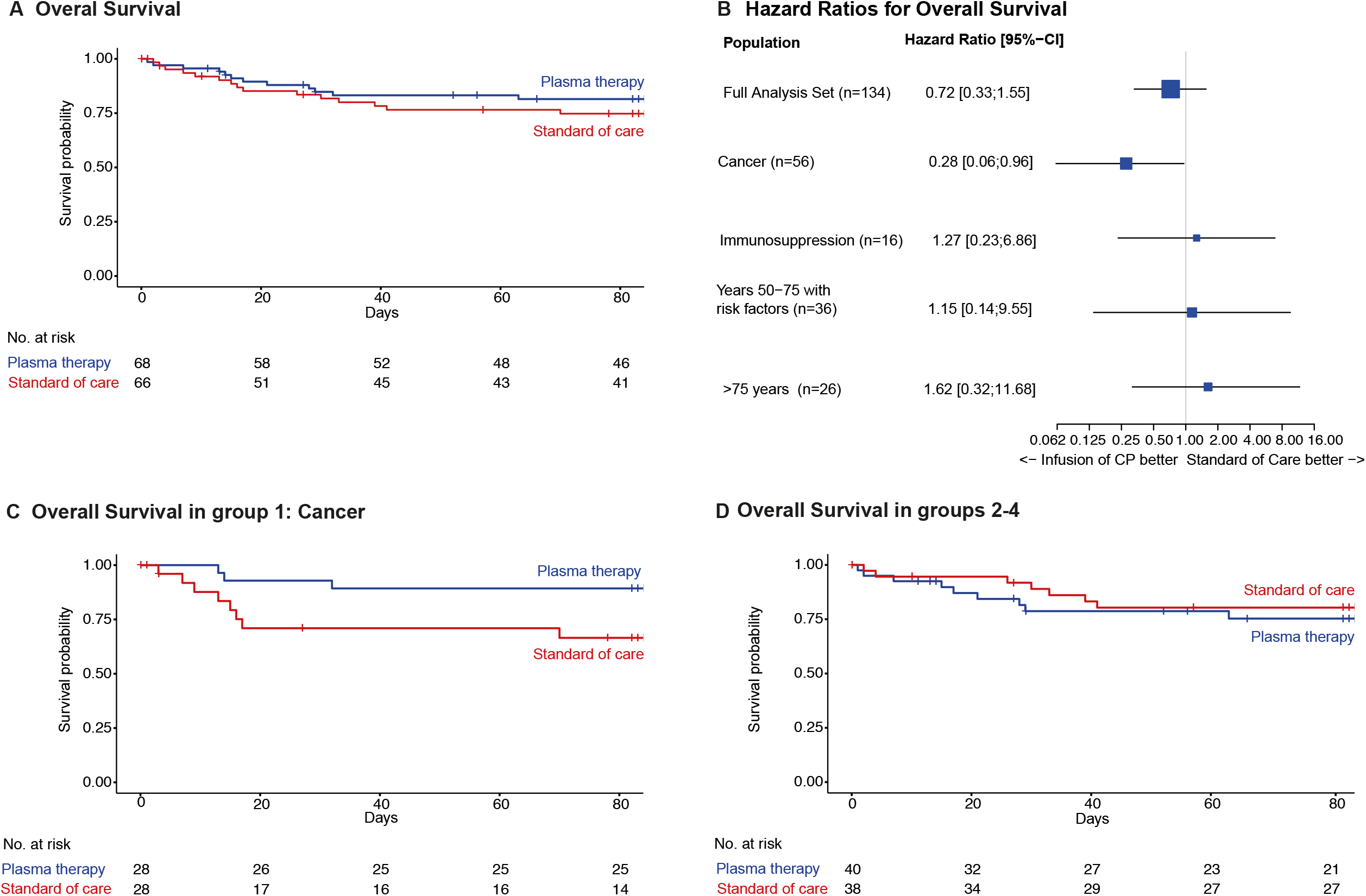
Secondary endpoint - Overall survival. A: Kaplan Meier curve for survival probability for overall study cohort (group-1 to -4) by PLASMA (blue) and CONTROL (red) with number of subjects at risk; log-rank *p*=0.403. B: Forest plot with hazard ratios (HR) for survival probability overall and by predefined subgroups, 95% confidence intervals in brackets. C: Kaplan Meier curve for survival probability for group-1 by PLASMA (blue) and CONTROL (red) with number of subjects at risk; log-rank *p*=0.042. D: Kaplan Meier curve for survival probability for combined group-2 to -4 by PLASMA (blue) and CONTROL (red) with number of subjects at risk. See Supplementary Figure S4 for separate data of groups 2-4; log-rank *p*=0.555.

Time to discharge did not differ (HR 1.28; 95%-CI 0.86-1.91; log-rank *p*=0.217) in the overall study population (PLASMA 12.5 days [95%-CI 10-17] versus CONTROL 18 days [95%-CI 11-28]) (Figure S4). Discharge occurred earlier in group-1 upon PLASMA (median 13 days; 95%-CI, 8-14) versus CONTROL (median 31 days ;95%-CI, 15-NA), (HR, 2.50; 95%-CI, 1.34-4.78; log-rank *p*=0.003).

Mechanical ventilation was initiated in 28.5% of patients. No significant difference was observed between the treatment groups (PLASMA 27.9% (95%-CI 18.7-39.6) versus CONTROL 29% (95%-CI 19.2-41.3), odds ratio (OR) 0.95 (95%-CI 0.44-2.06), *p*=0.892) or within the subgroups (Table 2). The outcome for patients that crossed over was not substantially different to other patients in the control arm.

### Neutralizing Antibody Titers

At the time of randomization, the average percent inhibition of SARS-CoV-2 virus measured with the surrogate neutralizing ELISA assay was 9.3% (IQR 4.8, 26.2; PLASMA 10.2, IQR 5.5, 28.8 versus CONTROL 8.5%, IQR 4.0, 20.3) (Figure 4A, Table S6). Neutralizing activity increased over time in both arms (Figures 4B, 4C, S6). Highest levels at day 3/5 were overall higher in the PLASMA cohort (PLASMA 51.1 [IQR 14.7, 92.5] compared to CONTROL 21.6 [IQR 7.2, 87.3]) (Figures 4B, 4C]). In patients with cancer, neutralizing activity did not increase over time in the absence of plasma therapy. In contrast, plasma therapy increased neutralizing activity in cancer patients with higher levels on day 3/5 (group-1; PLASMA 30.9, IQR 15.4, 98.0 compared to CONTROL 8.8, IQR 3.5, 46.3; Figures 4C, S5). Accordingly, for group-1 the median difference from day 3/5 to baseline differed significantly in PLASMA (9.1; IQR: 3.8, 24.9) compared to CONTROL (1.6, IQR: -1.5, 4.7; *p*=0.001, Figure 4C, left). In groups 3 and 4, neutralizing antibodies were already present at the time of study inclusion (Fig. S5) and titers further increased over time regardless of the therapy arm. Thus, there was no benefit in neutralizing antibody titers for group3 and 4 patients treated with plasma. Of note, in the few patients included in group 2 (immunosuppression), titers of neutralizing antibodies were low at the time of inclusion and remained low regardless of therapy arm (Fig. S5).

**Figure 4:**
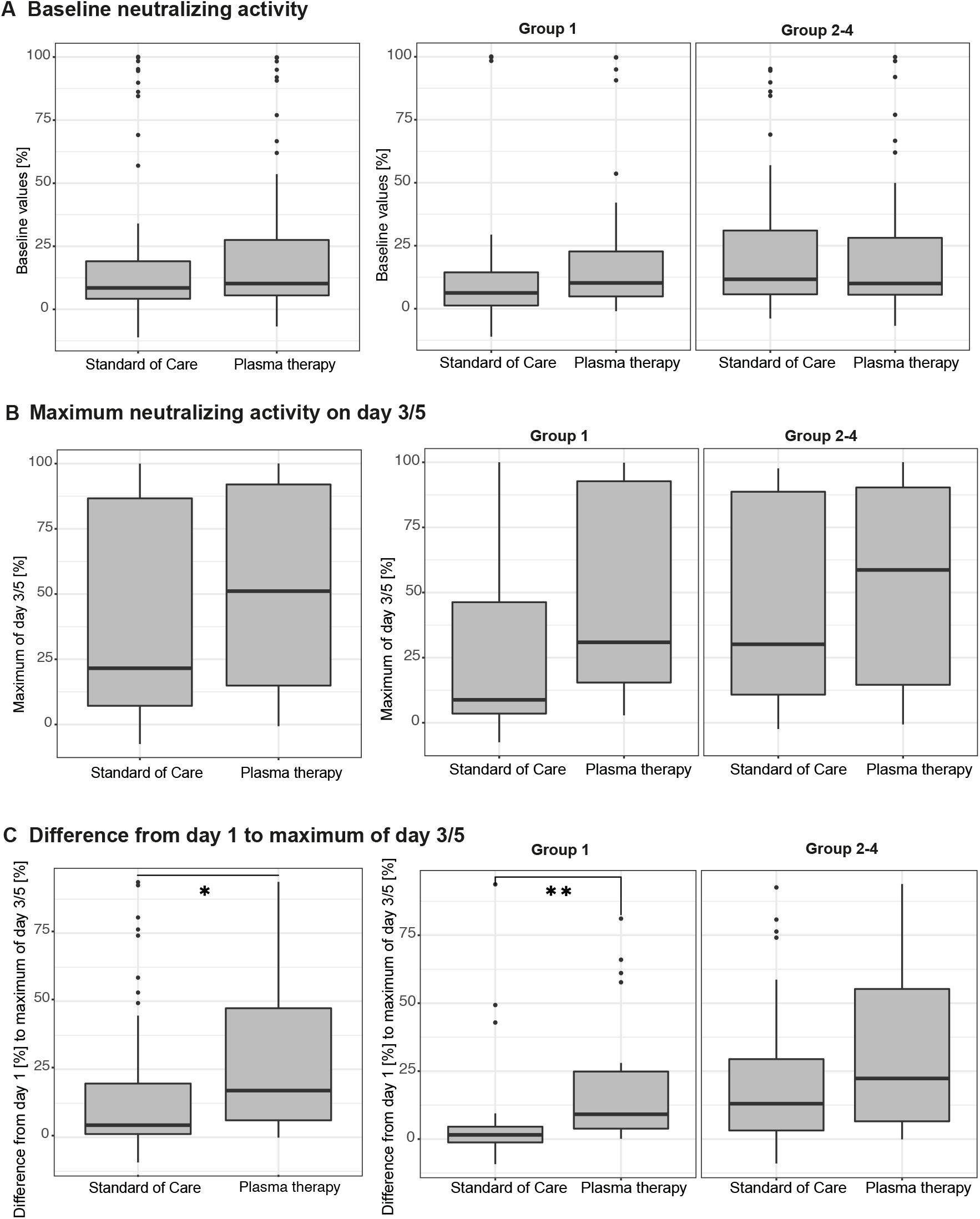
SARS-CoV-2 neutralizing activity in patient plasma. (A) Baseline neutralizing activity in the overall analysis set and group-1 and group-2 to -4 measured by a surrogate inhibition assay on day 1 (after randomization and prior to plasma treatment). Boxplots indicate the interquartile range and whisker length is limited to 1.5 times the interquartile range. Medians are indicated within the boxes. (B) Highest levels of neutralizing activity on day 3/5 in the overall analysis set and group-1 and group-2 to -4 measured by a surrogate inhibition assay. Boxplots indicate the interquartile range and whisker length is limited to 1.5 times the interquartile range. Medians are indicated within the boxes. (C) Increase in neutralizing activity analyzed as the percent difference in neutralizing activity as measured by a surrogate inhibition assay on day 1 (after randomization and prior to plasma treatment) and compared to the highest level of day 3/5 in the (A) overall analysis set and group-1 and group-2 to -4. Boxplots indicate the interquartile range and whisker length is limited to 1.5 times the interquartile range. Medians are indicated within the boxes. *\*p*=0.012, van-Elteren test stratified for patient group; *\*\*p*=0.001, Wilcoxon signed rank test.

### Adverse Events

Adverse events observed after plasma administration were in accordance with published data^17^. No serious adverse events were observed related to plasma therapy. Adverse events are provided in Table S7. Infusion reactions are described in Table S8.

## Discussion

Results of the RECOVER trial provide evidence that patients with cancer (group-1) who develop severe COVID-19 benefit from anti-SARS-CoV-2 plasma from convalescent/vaccinated donors and experience improved overall recovery. Although the size of group-1 was relatively small with 56 patients, differences in the primary endpoint were substantial (13 vs. 31 days) and are supported by earlier discharge and improved overall survival. The likelihood of improved outcomes upon plasma therapy was substantial for patients with cancer with shortened time to the primary endpoint, time to discharge and also survival. In contrast, no benefits were observed in groups-2 to -4 pointing towards a specific benefit of vaccinated/convalescent plasma in cancer patients. These results from a specifically designed clinical trial are in line with two retrospective propensity-matched cohort analyses, with in total 244 patients treated with plasma^27,28^ and one prospective non-randomized study using neutralizing monoclonal antibodies^14^.

Antivirals and monoclonal antibodies for COVID-19 are most effective in early disease-stages and are usually not recommended beyond 5 days after symptom-onset. The same was shown for early convalescent plasma therapy in a pre-hospital setting without need for supplemental oxygen^32^. In our study, patients were randomized and treated on average within 7 days of symptom-onset and few crossed-over at day 10 after randomization. An even earlier intervention with plasma might further increase efficacy in patients with cancer. Few therapies with proven efficacy are available for these patients at later time points, particularly for those presenting with impaired oxygenation, making plasma an attractive treatment approach even at advanced disease stages.

Unlike monoclonal antibodies^33^ convalescent plasma holds the potential to evolve in real-time with the virus and retain activity against new variants. Furthermore, it does not involve patent fees and can be obtained within the regular blood donor pool. Since plasma from vaccinated donors contained higher levels of neutralizing antibodies^34^, and antibody titer has been associated with efficacy^35^, we included vaccinated donors once vaccination was widely available (see Supplement Table S4).

Our study has limitations. Compared to studies with non-selected patients, the overall cohort is relatively small. Also, while the overall cohort was well balanced, we did observe imbalances between enrollment arms (e.g. in respect to age, sex, co-morbidities, therapy) in the subgroups. Therefore, we adjusted for the two variables (i.e. age and sex) most likely to be associated with outcomes in a sensitivity analysis, which did not impact on the primary outcome in group-1. Another limitation could be the open-label design of our trial. However, the results of subgroups of the primary endpoint are supported by results in the secondary endpoints overall survival and neutralizing activity showing the unique effectivity of plasma therapy in cancer patients.

The group of cancer patients was diverse with most patients suffering from hematological malignancies. Thus, the conclusions might not be applicable to all types of cancer. Lastly, recruitment occurred over an extended time span with different virus variants and evolving standards of care. Nonetheless, randomization was in place and plasma was obtained during the respective waves of the pandemic. Our conclusions formally cannot be extended to novel variants not covered within the trial (starting with Omicron). While studies have suggested that neutralizing antibodies were broadly active prior to Omicron, the Omicron variant in particular has demonstrated that variant-specific plasma would be important to control virus replication^33^.

There are several strengths of our trial: Plasma was obtained from donors with confirmed high titers of neutralizing antibodies as indicated by the fact that <20% of patients in the donor pool met the criteria (>= 1:80 titer and corresponding high saturation in the NeutraLISA; Figure S7). The relevant subgroups were pre-defined in the protocol. Hazard ratios and confidence intervals indicated large effect sizes in group-1. Plasma therapy effects on neutralizing antibody levels matched clinical benefit, although causality cannot be proven. The inclusion of patient groups now known not to benefit from plasma (e.g. groups-3 to -4) suggests that underlying disease characteristics determine the benefit of plasma therapy in cancer patients.

We found that in cancer patients an increase in neutralizing antibodies was observed after plasma infusion, which further supports the restriction of the beneficial effects of plasma to patients with limited ability to react to the antigen with a humoral response.

Taken together, these data suggest that plasma therapy may improve outcome in cancer patients with severe COVID-19.

## Methods

### Study sites and trial eligibility

Fifteen trial sites in Germany enrolled study participants (10 university and 5 urban hospitals, table S1 and S3). Adult patients requiring hospital admission for COVID-19 were assessed for eligibility irrespective of previous SARS-CoV-2 infection or vaccination status. Inclusion criteria were: (1) polymerase chain reaction (PCR) confirmed infection with SARS-CoV-2 in a respiratory tract sample; (2) oxygen saturation (SaO2) on ambient air of ≤94% or a partial oxygen pressure (PaO2) – inspired oxygen fraction (FiO2) ratio of <300mmHg; (3) provision of written informed consent; (4) meeting at least one high-risk criterion to define the patient group:

⍰ Group-1 – ‘cancer’: patients with pre-existing or concurrent hematological cancer and/or active cancer therapy for any cancer (incl. chemo-, radio- and surgical treatments) within the past 24 months.
⍰ Group-2 – ‘immunosuppression’: patients under chronic immunosuppression either pharmacological and/or due to underlying diseases not meeting Group-1 criteria.
⍰ Group-3 – ‘lymphopenia/elevated D-dimers’: patients aged >50 and ≤75 years not meeting Group-1 or -2 criteria who had lymphopenia (<0.8 G/l) and/or D-dimers (>1µg/ml).
⍰ Group-4 – ‘age >75 years’: patients aged >75 years not meeting Group-1, -2 or -3 criteria. Inclusion criteria were consecutively checked for groups-1 to -4 in ascending order.

Patients with a history of reaction to blood products, patients requiring mechanical ventilation (incl. non-invasive ventilation), selective IgA-deficiency and patients participating in another trial of investigational medicinal products were excluded. Further details on inclusion/exclusion are listed in the published protocol^31^. Modifications of the protocol and the statistical analysis plan are described in Table S2.

### Randomization and Intervention

Eligible patients underwent randomization into the experimental (PLASMA) or control (CONTROL) arm at a 1:1 ratio using block randomization for the patient group strata defined above (groups-1 to -4). Patients randomized into CONTROL were offered to cross over on day 10 (+ max. 2 days) after randomization in the absence of clinical improvement. CONTROL patients received standard of care as defined by the respective hospital at the time of trial inclusion. Patients in PLASMA received two units of AB0-compatible plasma, 238-337 ml each, from two different donors on the day of randomization (=day 1) and the day thereafter intravenously in addition to standard of care. Convalescent and/or vaccinated donor plasma was obtained at the IKTZ Heidelberg, Heidelberg, Germany. Plasma donor eligibility required high titers of neutralizing antibody activity in a live virus neutralization assay (titers ≥1:80; less than 20% of potential donors) (Supplementary text S2, S3, Figure S6, Table S4).

### Procedures

After obtaining informed consent, a clinical seven point ordinal scale (7POS)^29,30^ was determined daily, which was defined as: 1, not hospitalized with resumption of normal activities; 2, not hospitalized, but unable to resume normal activities; 3, hospitalized, not requiring supplemental oxygen; 4, hospitalized, requiring supplemental oxygen; 5, hospitalized, requiring nasal high-flow oxygen therapy, noninvasive mechanical ventilation, or both; 6, hospitalized, requiring ECMO, invasive mechanical ventilation, or both; and 7, death^31^.

### Endpoints

The primary endpoint was defined as time from randomization to a 2-point improvement on the 7POS or live hospital discharge, whichever occurred first. Secondary endpoints were overall survival (time from randomization until death from any cause); antibody titers; requirement of mechanical ventilation at any time during the hospital stay; time from randomization until live hospital discharge.

### Safety Assessments

All adverse events were graded according to Common Terminology Criteria for Adverse Events (CTCAE) version 5.0. Pharmacovigilance was performed according to the International Conference on Harmonization and Good Clinical Practice guidelines (ICH-GCP E6 (R2)). An independent data monitoring committee regularly assessed outcomes and Serious Adverse Events.

### Laboratory Analyses

Standard laboratory tests were performed locally. RT-PCR from nose/throat swabs for SARS-CoV-2 and antibody determination were performed in the Department of Infectious Diseases, Virology, Heidelberg. NeutraLISA assay (Euroimmun, Lübeck, Germany) measures serum competition with ACE2-S1 binding and was used as a surrogate for neutralizing SARS-CoV-2 antibody activity in plasma. Live virus neutralization assay and NeutraLISA correlation for donor plasma is provided in Figure S7.

### Early trial termination

The first patient was randomized on September 3rd, 2020. Enrolment fluctuated with SARS-CoV-2 incidence in Germany. In January 2022 the omicron variant became dominant in Germany. Neutralizing activity of stored plasma against omicron was unknown. Also, enrolment had slowed considerably after new guidelines (from World Health Organization and others) on convalescent plasma use. The Data Monitoring Board thus recommended to stop recruitment, which was enacted on January 20th, 2022, after enrolment of 77% of the target population.

### Statistical analysis

The analysis of the primary endpoint was done via a log-rank test, stratified for the factor “patient group”. The event “death from any cause” was handled by censoring those patients at day 84^36^. Hazard ratios were determined via Cox regression stratified by patient group (1-4). A post-hoc sensitivity analysis was performed using an adjusted Cox regression considering age and sex to account for differences observed in the distribution of these variables between study arms. Time to discharge was assessed analogously to the primary endpoint. Overall survival was assessed by means of a log-rank test and Cox regression, both stratified for patient group. Requirement of mechanical ventilation (yes/no) was analyzed by means of a logistic regression model adjusting for the factors treatment and patient group including all patients with more than 1 day of follow-up. Patients who died were accounted for as having received mechanical ventilation. For neutralizing antibodies, the difference between baseline and the highest value on day 3/5 was assessed to compare the PLASMA vs. CONTROL titers stratified by patient group (1-4) and a van Elteren test was performed. Predefined subgroup analyses were conducted for each patient group, as well as an exploratory analysis of the treatment effect interaction between the patients in group-1 versus groups-2 to -4 combined. Patients with incomplete follow-up after discharge were censored at the date of last follow-up. Complete case analyses were performed, and no imputation of missing data was conducted. Adverse events were summarized descriptively. The analysis of efficacy endpoints was done in the full analysis set including all randomized patients, while the safety endpoints were analyzed according to the treatment actually received. The trial was designed to enroll 174 patients (87 per arm) to detect a hazard ratio of 1.6 for shortening the time to improvement of 2 points on the 7-point ordinal scale or live hospital discharge in PLASMA compared to CONTROL at a two-sided significance level of 5% with a power of 80%. Additional details are provided in the protocol and statistical analysis plan (text supplement S1 and S4). The statistical analysis plan was written while investigators were blinded to treatment allocation.

### Ethics and regulatory

This study was carried out in accordance with the Declaration of Helsinki and the ICH-GCP E6 (R2) guidelines. The study was approved by the federal institute for vaccines and biomedicines (Paul-Ehrlich-Institute) and the ethics committee Heidelberg. Regulatory authority requirements with respect to plasma manufacturing according to §67 Arzneimittelgesetz (Germany) and §13 GCP-V were met.

### Trial registration

EudraCT Number: **2020-001632-10**, https://www.clinicaltrialsregister.eu/ctr-search/trial/2020-001632-10/DE, registered on 04/04/2020.

## Data Availability

All data produced in the present study are available upon reasonable request to the authors

## Funding

The trial was co-financed by the Federal Ministry of Education and Research, Germany (BMBF; emergency research funding RECOVER 01KI20152).

## Data availability

Please contact for data availability.

## Code availability

The full code will be accessible upon publication within a git. Please contact for code availability.

## Notes

### Competing Interest Statement

The authors have declared no competing interest.

### Clinical Trial

EudraCT Number 2020-001632-10

### Author Declarations

Ethics committee of University of Heidelberg gave ethical approval for this work

