## Supplement for "Anti-SARS-CoV-2 antibody containing plasma improves outcome in patients with hematologic or solid cancer and severe COVID-19 via increased neutralizing antibody activity – a randomized clinical trial"

Claudia Denkinger, Maike Janssen et al.,

Table of contents

1. Texts
   - Text supplement S1 Study protocol
   - Text supplement S2 Information on Plasma donation
   - Text supplement S3 Neutralization assay used for determination of titer in donors
   - Text supplement S4 Statistical analysis plan
2. Figures
   - Figure S1 Consort diagram for group-1 (hematological and solid cancer)
   - Figure S2 Kaplan Meier curves for time to improvement on 7-point ordinal scale or live hospital discharge for group-2 to -4
   - Figure S3 Kaplan Meier curves for Survival for group-2 to -4
   - Figure S4 Kaplan Meier curves for Time to discharge overall
   - Figure S5 NeutraLISA measurement in the four subgroups
   - Figure S6 Column scatter plots for neutralization characteristics of donated plasma
   - Figure S7 Correlation between live virus neutralization assay and NeutraLISA in donor plasma
3. Tables
   - Table S1 Trial sites, enrolled patients and local investigators
   - Table S2 Protocol amendments and SAP deviations
   - Table S3 Representativeness
   - Table S4 Plasma donor characteristics
   - Table S5 Epidemiological data of study participants in predefined subgroups
   - Table S6 Baseline values and treatment in predefined subgroups
   - Table S7 Adverse Events
   - Table S8 Infusion reactions
1. **Text supplements**

Text supplement S1 Study protocol

Previously published: [1] and separately provided

Text supplement S2 Information on Plasma donation

Convalescent plasma was procured from individuals after a minimum of three weeks from symptom resolution throughout the duration of the trial. Plasma from vaccinated individuals (used since May 18, 2021) was obtained from donors with at least two doses of mRNA vaccines. Only plasma from consented donors with high levels of neutralizing antibodies against the prevailing variant at the time (titers ≥ 1:80; less than 25% of donors) at initial evaluation was used (details on the neutralizing assay are provided below). Plasma was later also characterized with NeutraLISA assay (Euroimmun, Lübeck, Germany) for comparability against measurements in patients. Plasma from COVID-19 convalescent patients or vaccinated healthy donors (CVP) was collected according to standard operating procedures for fresh-frozen plasma in accordance with national regulatory requirements and EU- Good Manufacturing Practice (GMP) Guidelines. A specific manufacturing license for plasma within the RECOVER trial was obtained from the regional authorities (Regierungspraesidium Tuebingen). Plasma was frozen and shipped to trial sites upon randomization of a patient into the plasma-arm of the study.

Text supplement S3 Live virus neutralization assay used for determination of neutralizing antibody titer in donors

VeroE6 cells were seeded into 96-well plates one day before the assay. Serial dilutions of sera starting from a 1:10 dilution were prepared in OptiMEM (ThermoFisher Scientific, Waltham, MA, USA) in a final volume of 75 µl and incubated with ~24,000 plaque-forming-units of SARS-CoV-2 (BavPat1/2020 strain, European Virus Archive) per well for 1 hour at 37 °C in a final volume of 150 µl. One third of each serum-virus mixture (50 µl) was then used for infection (final multiplicity of infection ~0.25). Infections were performed in duplicates. At 20 hours post-infection, cells were fixed with 5% formaldehyde and immunostained using a primary mouse antiserum binding to double-stranded RNA, a viral replication intermediate (J2 mouse antibody; Scicons, Szirák, Hungary) and a secondary anti-mouse horseradish peroxidase-coupled antibody (Merck, Darmstadt, Germany). The signal was developed using the KPL SureBlueTM TMB peroxidase substrate (Seracare, Milford, MA, USA) and measured in a plate reader at a wavelength of 450 nm. Data were normalized to no-serum control (100%) and mock-infected control (0%). The neutralization (NT) titer was defined as the highest serum dilution resulting in more than 50% reduction of the normalized signal. Testing was performed against the most prevalent variant at the time (wildtype, alpha, delta).

Text supplement S4 Statistical analysis plan

**SAP for Final Analysis**

**RECOVER**

**A Randomized Open label Phase-II Clinical Trial with or without Infusion of Plasma from Subjects after Convalescence of SARS-CoV-2 Infection in High-Risk Patients with Confirmed Severe SARS-CoV-2 Disease**

**Clinical Trial Code: SARS-CoV-2CP-HD-001**

**EudraCT No.: 2020-001632-10**

**SAP refers to Protocol version 3.0**

**1 Purpose of this SAP**

In the trial protocol the assumptions, methods and procedures for the statistical evaluation are already described. The statistical analysis plan (SAP) aims to specify more precisely the evaluations carried out in the statistical analysis. This SAP refers to the final analysis after study completion, no interim analysis was planned or conducted.

**2 Objective of the Trial**

The primary objective of the trial is to assess the time from randomization until an improvement within 84 days defined as two points on a seven point ordinal scale (given in chapter 5.1) or live discharge from the hospital in high-risk patients (group 1 to group 4 as defined below) with SARS-CoV-2 infection requiring hospital admission by infusion of plasma from subjects after convalescence of SARS-CoV-2 infection/vaccination against SARS-CoV-2 or standard of care.

The four high-risk patient groups are:

1. Patients with pre-existing or concurrent hematological malignancy and/or active cancer therapy (including chemotherapy, radiotherapy, surgery) within the last 24 months or less.
2. Patients with chronic immunosuppression not meeting the criteria of group 1.
3. Patients of age ≥ 50 - 75 years meeting neither the criteria of group 1 nor group 2 and at least one of these criteria: Lymphopenia < 0.8 x G/l and / or D-dimer > 1μg/mL.
4. Patients of age ≥ 75 years meeting neither the criteria of group 1 nor group 2.

The link between the primary objective and the primary analysis is also described in the sense of the new estimands framework which is given in chapter 10.

Secondary objectives of the trial are:

- to assess overall survival, and the overall survival rate at 28, 56 and 84 days,
- to assess SARS-CoV-2 viral clearance and load as well as antibody titres,
- to assess percentage of patients that required mechanical ventilation, and
- to assess time from randomization until discharge
- to assess cytokine changes over time.

**3 Study design**

RECOVER is a randomized, open-label, multicenter phase II trial, designed to assess the clinical outcome of SARS-CoV-2 disease in high-risk patients (group 1 to group 4) following treatment with plasma from subjects after convalescence of a SARS-CoV-2 infection or after vaccination against SARS-CoV-2 compared with standard of care. Randomisation is done either into the experimental arm “infusion of convalescent plasma (CP) or vaccine-boosted plasma (=Post-Vaccination Plasma/PVP) ” or into the standard arm “standard of care” in a 1:1 ratio using block randomisation stratified for the factor “patient group”, defined by groups 1 to 4 as given in Chapter 2. The total planned sample size is n = 174 (84 per group).

**4 Analysis sets**

Full Analysis Set

The Full Analysis Set (FAS) includes all randomised patients with treatment groups assigned in accordance with the randomisation, regardless of the treatment actually received. Patients who were randomised but did not subsequently receive treatment are included in the FAS. The analysis of data using the FAS follows the Intention-to-Treat principles (ITT).

Per Protocol Set

In the per protocol set (PPS), patients with important protocol deviations (e.g. patients not meeting the inclusion/exclusion criteria, too early cross-over, reception of only one instead of two bags of infusion) are excluded. Analyses based on the PPS are known to be biased and are therefore of minor importance for the interpretation of the results and only serve as supplementary analyses. The determination of patients with major protocol deviations will be done in a data review meeting which will take place before database lock.

Safety Set

All enrolled patients who received treatment are subjected to the safety analysis. Patients will be evaluated with regard to the treatment actually received, meaning that patients who cross over from the control to the experimental arm will be evaluated in the experimental arm at day 10 (+ 2 days) onwards.

**5 Definitions of endpoints to be analysed**

**5.1 Primary Endpoint**

The primary efficacy endpoint is time from randomisation to clinical improvement within 84 days by two points on a seven point ordinal scale (in comparison to the status at randomisation) or live discharge from the hospital. If a patient needs to be transferred to another hospital, he or she will not be considered as discharged. If a patient is transferred to a rehabilitation clinic or nursing home, he or she will be considered as discharged. The categories of the seven point ordinal scale are:

1. Not hospitalized with resumption of normal activities
2. Not hospitalized, but unable to resume normal activities
3. Hospitalized, not requiring supplemental oxygen
4. Hospitalized, requiring supplemental oxygen
5. Hospitalized, requiring nasal high-flow oxygen therapy, or noninvasive mechanical ventilation
6. Hospitalized, requiring ECMO, invasive mechanical ventilation, or both
7. Death

**5.2 Secondary Endpoints**

Secondary endpoints are:

- Overall survival, defined as the time from randomization until death from any cause
- 28-day, 56-day and 84-day overall survival rate
- Requirement of mechanical ventilation at any time during hospital stay (yes/no)
- Time from randomization until discharge. If a patient needs to be transferred to another hospital, he or she will not be considered as discharged. If a patient is transferred to a rehabilitation clinic or nursing home, he or she will be considered as discharged.
- SARS-CoV-2 viral clearance and load as well as antibody titres (until hospital discharge on day 1, 3, 5, 10, 14, 28, 56 and 84), comprising:
  - Result of smear (negative / positive / invalide)
  - CT-value
  - Serodiagnostic by ELISA (OD ratio)
  - Neutralizing antibody titre
- Cytokine profiles

**5.3 Exploratory Endpoints**

Exploratory endpoints are:

- Vital signs (on day 1, 3, 5, 10 and 14), comprising:
  - ECOG performance status (ordinal, 0 to 4)
  - Systolic blood pressure in mmHg
  - Diastolic blood pressure in mmHg
  - Pulse rate in beats/min
  - Respiratory rate in breaths/min
- Seven point ordinal scale as defined in chapter 5.1 (until hospital discharge daily from day 1-28, weekly from day 29-55 and on day 56 and 84)
- Sequential Organ Failure Assessment (SOFA) Score (until hospital discharge on day 1 and 10)
- Oxygen saturation:
  - Partial pressure of oxygen (PaO_2_) in mmHg
  - Partial pressure of carbon dioxide (PaCO_2_) in mmHg
  - ECMO (yes / no)
  - Type of ventilation (no / nasal high-flow oxygen therapy / noninvasive mechanical ventilation / invasive ventilation / tracheotomy / ambient air / standard oxygen sources)
  - Amount of oxygen in litres/min
  - Oxygen saturation (SaO_2_) in percent
  - Amount of supplemental oxygen that is required to keep SaO_2_ above 94% in litres/min
  - Fraction of inspired oxygen (FiO_2_) in percent
- Hematology (until hospital discharge on day 1, 3, 5, 10, 14, 28, 56 and 84), comprising:
  - Hemoglobin
  - RBC
  - WBC
  - Thrombocytes
- Blood chemistry and coagulation (until hospital discharge on day 1, 3, 5, 10, 14, 28, 56 and 84), comprising:
  - BUN
  - Creatinine
  - Albumin
  - AST/SGOT
  - ALT/SGPT
  - Total bilirubin
  - GGT
  - AP
  - LDH
  - Sodium
  - Potassium
  - Magnesium
  - Calcium
  - Uric acid
  - Troponin
  - CK
  - CK-MB
  - PTT
  - ATIII
  - D-dimer
  - Fibrinogen
  - Ferritin (measured at day 1 only)
  - Transferrin (measured at day 1 only)
  - Transferrin saturation (measured at day 1 only)
  - CRP
  - Total protein (measured at day 1 only)
  - IL6 (measured at day 1 only)
  - Procalcitonin (measured at day 1 only)
  - Total IgG (measured at day 1 only)
  - IgA (measured at day 1 only)
  - IgM (measured at day 1 only)
  - Lactate
  - INR
- Assessment of concomitant medications
- Signs/symptoms (on day 10 in case of cross-over), comprising:
  - Cough (yes / no)
  - Fever (yes / no)
  - Myalgia (yes / no)
  - Fatigue (yes / no)
  - Diarrhoea (yes / no)
  - Vomiting (yes / no)
  - Headache (yes / no)
  - Abdominal pain (yes / no)
  - Nausea (yes / no)
  - Olfactory or taste disorder (yes / no)
  - Dyspnoea (yes / no)
  - Confusion (yes / no)
- Physical examination (on day 10 in case of cross-over), comprising:
  - General appearance (normal / abnormal)
  - Lung (normal / abnormal)
  - Cardiac auscultation (normal / abnormal)
  - Cardiac percussion (normal / abnormal)
  - Abdominal auscultation (normal / abnormal)
  - Abdominal percussion (normal / abnormal)
  - Abdominal palpation (normal / abnormal)
  - Palpation of lymph node sites (normal / abnormal)
  - Neurological examination (normal / abnormal)
  - Peripheral edema (normal / abnormal)
- ECG, comprising
  - ECG abnormal / abnormal
  - Clinically significant (yes / no)
  - Heart rate in beats/min
  - Rhythm (normal sinus / other)

**5.4 Safety Endpoints**

Safety endpoints comprise:

- Adverse Events (AEs)
- Severity of AEs
- Serious adverse events (SAEs)
- Relation of AEs to the study treatment
- Dose modifications for toxicity
- Discontinuation of study treatment

**6 Methods for Withdrawals and Missing Data**

Withdrawals according to study protocol will not be excluded unless the patient withdrew his/her consent declining explicitly the use of the data for analysis (s. informed consent). As long as a written informed consent is available without denied permission to evaluate the documented data, the data already collected as well as follow-up data will be included in the final analysis.

For patients with incomplete follow-up, time to last follow-up date is used as the censoring time in the analysis of time-to-event data. Otherwise, no imputation of missing data will be conducted.

**7 Statistical methods**

The statistical evaluation is carried out under the supervision of the Supervising Statistician. Statistical analysis is based on the International Conference on Harmonization (ICH) Guidelines “Structure and Content of Clinical Study Reports” and “Statistical Principles for Clinical Trials”. All statistical procedures are done according to the current Standard Operating Procedures (SOPs) of the Institute of Medical Biometry, University of Heidelberg (IMBI).

***7.1 Descriptive methods***

A CONSORT flow diagram will be created to display the progress of all participants through the trial. This includes the number of patients assessed for eligibility and the number of patients excluded because they did not meet inclusion criteria, declined to participate, or any other reason, the number of randomized patients (per group), and number of missed intervention assignments / incomplete interventions. In addition, the number of patients in the FAS and PPS will be given and reported reasons for exclusion from the PP set will be summarized per treatment group.

Continuous variables will be described using number of observations, mean, standard deviation, median, Q1, Q3, minimum, maximum and number of missing values. For categorical variables, absolute and relative frequencies will be given with missing values being reported as a separate category. Percentages for categorical variables will be based on all non-missing values in the respective groups. Time to event endpoints will be described by Kaplan-Maier estimates (including the numbers at risk for different time points), median survival time (or other quantiles if median survival time cannot be calculated), number of events and number of censorings. The descriptive methods described above will be used separately for each treatment group.

The test to be applied is specified in the description of the analyses of the respective endpoint. If a test is conducted to compare treatment groups, point estimates with respective 95% confidence intervals will be provided additionally to the descriptive p-value. For the chi-square test, the difference of rates and/or the risk ratio will be given where appropriate, for the t-test the difference between means will be provided and for the Mann-Whitney U test, the Hodges-Lehmann estimate will be presented as point estimate.

Descriptive results will be presented in tables, appropriately supplemented with figures if specified in the respective analysis section. Survival endpoints will always be supplemented with a Kaplan-Meier survival curve. Listings will be used to present data collected in free-text formats.

**7.2 Baseline characteristics**

The following baseline characteristics will be collected and described in the FAS set, the PP set and the safety set:

- Demographics, comprising:
  - Sex (male / female)
  - Year of birth
  - Ethnic origin (asian / black / caucasian/white / hispanic / other)
- Medical history
  - Haematological malignancy
  - Solid tumor
  - Immunosuppression due to disease (immunodeficiency)
  - Diabetes
  - Cardiovascular disease
  - Obesity
  - Lung disease
- – Assessment of Concomitant medications
  - COVID19 therapy from 7 days before study entry up to day 10
    - Steroids
    - Remdesivir or other antiviral drugs
    - Monoclonal antibodies
    - Tociluzimab
    - Ruxolitinib
- Smoking history, comprising
  - Yes / non-smoker / ex-smoker
  - Smoking duration in years
  - Smoking amount in packs per day
- Signs/symptoms, comprising:
  - Cough (yes / no)
  - Fever (yes / no)
  - Myalgia (yes / no)
  - Fatigue (yes / no)
  - Diarrhoea (yes / no)
  - Vomiting (yes / no)
  - Headache (yes / no)
  - Abdominal pain (yes / no)
  - Nausea (yes / no)
  - Olfactory or taste disorder (yes / no)
  - Dyspnoea (yes / no)
  - Confusion (yes / no)
- Vital signs, comprising:
  - ECOG performance status (ordinal, 0 to 4)
  - Body temperature in degrees Celsius
  - Systolic blood pressure in mmHg
  - Diastolic blood pressure in mmHg
  - Pulse rate in beats/min
  - Respiratory rate in breaths/min
- Physical examination, comprising:
  - General appearance (normal / abnormal)
  - Lung (normal / abnormal)
  - Cardiac auscultation (normal / abnormal)
  - Cardiac percussion (normal / abnormal)
  - Abdominal auscultation (normal / abnormal)
  - Abdominal percussion (normal / abnormal)
  - Abdominal palpation (normal / abnormal)
  - Palpation of lymph node sites (normal / abnormal)
  - Neurological examination (normal / abnormal)
  - Peripheral edema (normal / abnormal)
- Clinical Frailty Scale (very fit / well / managing well / vulnerable / mildly frail / moderately frail / severely frail / very severely frail / terminally ill)
- Seven point ordinal scale as defined in chapter 5.1
- Oxygen saturation
  - Partial pressure of oxygen (PaO_2_) in mmHg
  - Partial pressure of carbon dioxide (PaCO_2_) in mmHg
  - ECMO (yes / no)
  - Type of ventilation (no / nasal high-flow oxygen therapy / noninvasive mechanical ventilation / invasive ventilation / tracheotomy / ambient air / standard oxygen sources)
  - Amount of oxygen in litres/min (will only be evaluated for any time point if patient does not simultaneously receive any oxygen)
  - Oxygen saturation (SaO_2_) in percent (will only be evaluated for any time point if patient does not simultaneously receive any oxygen)
  - Amount of supplemental oxygen that is required to keep SaO_2_ above 94% in litres/min
  - Fraction of inspired oxygen (FiO_2_) in percent
- ECG, comprising
  - ECG abnormal / abnormal
  - Clinically significant (yes / no)
  - Heart rate in beats/min
  - Rhythm (normal sinus / other)
- Hematology (until hospital discharge on day 1, 3, 5, 10, 14, 28, 56 and 84), comprising:
  - Hemoglobin
  - RBC
  - WBC
  - Thrombocytes
- Blood chemistry and coagulation (until hospital discharge on day 1, 3, 5, 10, 14, 28, 56 and 84), comprising:
  - BUN
  - Creatinine
  - Albumin
  - AST/SGOT
  - ALT/SGPT
  - Total bilirubin
  - GGT
  - AP
  - LDH
  - Sodium
  - Potassium
  - Magnesium
  - Calcium
  - Uric acid
  - Troponin
  - CK
  - CK-MB
  - PTT
  - ATIII
  - D-dimer
  - Fibrinogen
  - Ferritin (measured at day 1 only)
  - Transferrin (measured at day 1 only)
  - Transferrin saturation (measured at day 1 only)
  - CRP
  - Total protein (measured at day 1 only)
  - IL6 (measured at day 1 only)
  - Procalcitonin (measured at day 1 only)
  - Total IgG (measured at day 1 only)
  - IgA (measured at day 1 only)
  - IgM (measured at day 1 only)
  - Lactate
  - INR
- Urinalysis, comprising:
  - pH-value
  - Glucose
  - Protein
- Other virology, comprising:
  - Anti-HAV (negative / positive)
  - Anti-HBV (negative / positive)
  - Anti-HCV IgG (negative / positive)
  - HIV (negative / positive)

Baseline characteristics will be summarized for each treatment group separately using the methods described in Section 7.1.

**7.3 Primary analysis**

The primary analysis will assess the null hypothesis “the cumulative improvement curves for the primary endpoint in the experimental and control arm are equal”, i.e.

H_0_: S_E_ = S_C_

against the alternative hypothesis

H_1_: S_E_ ≠ S_C_

at a two-sided significance level of α=5%. This will be achieved by using the log-rank test stratified for the factor “patient group” as used in the randomisation procedure. The event “death from any cause” will be handled by censoring those patients at day 84 (in analogy to the approach of Cao et al, NEJM 2020). Using this approach ensures that deceased patients are considered as “not improved” over the whole observation period of 84 days. The primary analysis will be based on the FAS and handles intercurrent events as described in chapter 11.1.

The main estimate (corresponding to the main estimand as described in chapter 11.1) for the hazard ratio for treatment group will be computed using a Cox regression model stratifying for the factor “patient group” together with a 95% profile-likelihood confidence interval. The cumulative improvement curves are calculated using the Kaplan-Meier method together with 95% log-log-type confidence bands, and will be calculated separately for both treatment groups. In addition, cumulative improvement curves will also be calculated separately for each patient group per treatment group, and descriptive log-rank tests will be done to compare the cumulative improvement groups for each patient group. Patients who drop out during the trial or are lost to follow-up are taken into account as censored observations.

**7.4 Sensitivity and supplementary analyses**

All sensitivity analyses are conducted in the FAS and handle intercurrent events as described in chapter 11.1. If a supplementary analysis is conducted in a different analysis set or handles an intercurrent event differently this is explicitly mentioned.

**7.4.1Sensitivity analysis 1**

An unstratified log-rank test will be conducted, which is based on the assumption of identical baseline risk in the different patient groups. As a sensitivity estimate a hazard ratio for the treatment group based on an unstratified Cox-regression will be calculated together with a 95% profile-likelihood confidence interval.

**7.4.2 Sensitivity analysis 2**

A Gray’s test (Gray, 1988) stratified for “patient group” will be conducted where the death from any cause without previous improvement is treated as a competing risk. As a sensitivity estimate a subdistribution hazard based on a stratified Fine and Gray’s competing risk regression using PROC PHREG will be calculated together with 95% confidence intervals.

**7.4.3 Supplementary analysis 1**

A supplementary analysis of the primary endpoint will involve the inverse probability censoring weighting (IPCW) approach to estimate a treatment effect in the hypothetical scenario that patients from the control group had not switched to the experimental group (Robins & Finkelstein (2000)). This type of model requires to estimate the probability for a patient to switch to the experimental group based on (time-dependent) covariates, which in our case will be the baseline factor “patient group”, together with the time-varying factors “seven point ordinal scale measuring clinical improvement” and “viral load”. For this analysis, patients who cross over to the standard treatment will be considered as censored observations. Thus, however, patients who crossover due to poor prognosis will not contribute to the same degree as patients with a favourable prognosis less likely to switch. Thus, in order to adjust for informative censoring in case of treatment switch, patients in the control group will receive a weighting, such that uncensored patients with a high crossover risk due to unfavourable prognostic factors will receive higher weights than patients with a low crossover risk due to favourable prognostic factors. It should be noted that this method relies on the assumption that there are no unmeasured confounders influencing the probability to crossover, which is hardly verifiable, results will therefore needed to be cautiously interpreted.

o conduct the IPCW method, firstly, the data needs to be transformed into a long format, and the follow-up period needs to be split up in time intervals. In a first step, the probability to remain on the assigned treatment will be estimated for the control group using a repeated measures logistic regression model via PROC GENMOD with an unstructured covariance matrix, including only the patient group and the baseline ordinal scale as predictors. This model will estimate the probabilities

$$p_{ij} :=P\left[ No crossover for patient i in follow-up period \left[ t_{j},t_{j+1} \right) \right| Baseline covariates]$$

will be calculated. Subsequently, the probability for each control group patient to remain on the assigned treatment will be estimated using additionally the time-varying seven-point ordinal scale and the viral load, and probabilities

$$p_{ij}^{*} :=P\left[ No crossover for patient i in follow-up period \left[ t_{j}{,t}_{j+1} \right) \right| Baseline and time-dependent covariates]$$

will be given. Then, stabilized weights for each time period $\left[ t_{j}{,t}_{j+1} \right)$, j=1,… for each patient i will be calculated by $w_{ij}:=\prod_{k=1}^{j} p_{ik}/\prod_{k=1}^{j} p_{ik}^{*}$. These are the so-called stabilized weights proposed by Robins et al. (2000), which ensure that the weights are not too extreme, i.e. not too far away from 1. The weights of crossover patients are set to zero after the crossover occurs. Patients from the treatment group will receive constant and equal weights of 1 since they do not crossover. Then, a cox proportional hazard model with the time-varying weights $w_{ij}$ and the fixed factor treatment arm stratified for patient group will be calculated to estimate an adjusted hazard ratio for the treatment effect for the hypothetical scenario that no switching occurred.

**7.4.4 Supplementary analysis 2**

As another supplementary analysis intending to estimate the treatment effect in the hypothetical scenario which assumes that control patients had not switched to the treatment group, a rank preserving structural failure time (RPSFT) model will be fitted (Robins & Tsiatis, 1991). This model assumes a common treatment effect of the intervention, meaning that the treatment effect is the same for all individuals (with respect to time spent on treatment) regardless of when treatment is received, which is a strong and hardly verifiable assumption, thus requiring cautious interpretation of the modelled treatment effect.

As described by Latimer et al. (2017), the method splits the observed event time, $T_{i}$, for each patient into two, that is the event time when the patient is on the control treatment, $T_{A_{i}}$ , and the event time when the patient is on the intervention treatment, $T_{B_{i}}$. For patients who are randomised to the intervention treatment, $T_{A_{i}}$ is equal to zero. For patients randomised to the control group who do not switch onto the intervention (i.e. compliance is full in the control group), $T_{B_{i}}$. is equal to zero. However, for patients who switch treatments, both $T_{A_{i}}$ and $T_{B_{i}}$ will be greater than zero. The RPSFT method relates $T_{i}$ to the counterfactual event time ($U_{i}$), i.e. the time an individual would have had, had he or she received the control treatment, with the following causal model: $U_{i}=T_{A_{i}}+e^{\psi_{0}}T_{B_{i}}$. The value of $\psi$is estimated using a grid search, and the true value of $\psi=\psi_{0}$ is that for which $U_{i}\left( \psi\right)$ is independent of the randomised group. This will be assessed via a logrank test stratified for patient group. $\psi$ will be determined using the approach by Danner & Sarkar (2018), and event times for the control group adjusted via this counterfactual model (i.e. a comparison of ${e^{{-\psi}_{0}}U}_{i}$for patients in the intervention arm with $U_{i}$ for patients in the control arm) will be assessed via Kaplan Meier Curves and a Cox regression model stratified for patient group, producing an adjusted hazard ratio.

This adjustment also needs to be considered when determining event status for crossover patients on the control arm, as the recovery time for patients who crossed over from the control arm is likely to be artificially prolonged via the RPSFT approach, leading to fewer events in the control arm due to the data cutoff at 84 days. A correction to minimize this effect in censoring time is mentioned in Robins and Tsiatis (1991), and they propose to use Min (C, C*exp($\psi$)) as the date of administrative censoring, where C is the follow-up time for each patient. As $\psi$ is expected to be positive for the primary endpoint of the RECOVER trial due to the expected benefit in recovery time, we will use Max(C, C*exp($\psi$)) as the date of administrative censoring, thereby increasing the “hypothetical” follow-up time for the control arm. It should be noted that for the RPSFT analyses on overall survival, the originally proposed adjustment of Min (C, C*exp($\psi$)) will be done.

**7.4.5 Supplementary analysis 3**

As a further supplementary analysis the analysis as described in chapter 7.3 will be conducted in the PPS as defined in chapter 4. A supplementary estimate will also be calculated based on the Cox regression model described in chapter 7.3. Analyses in the PPS don’t target any causal effect and are known to be biased, hence results of these supplementary analyses are of secondary importance to the interpretation of the trial results.

**7.4.6 Supplementary analysis 4**

As a further supplementary analysis the analysis as described in chapter 7.3 will be conducted in the as-treated set, in which patients who crossed over will be evaluated in the experimental group. A supplementary estimate will also be calculated based on the Cox regression model described in chapter 7.3.

**7.4.7 Supplementary analysis 5**

A further supplementary analysis will focus on the effect of cross-over. To evaluate the effect of cross-over of patients randomized into the Standard-treatment arm, a subset of patients eligible for cross-over is defined (still hospitalized at day 10, no improvement on the 7 point ordinal scale) and analyzed with regard to the primary and secondary endpoints according to plasma-treatment.

**7.5 Further analyses based on the primary endpoint**

**7.5.1 Impact of timing of treatment**

To assess the impact of the timing of treatment, a Cox regression model adjusting for the factor treatment and time between randomization and first dose of treatment, stratified for patient group, is conducted. The hazard ratio and a 95% profile-likelihood confidence interval for “time between randomization and first dose of treatment” will be obtained from the model.

**7.5.2 Impact of type of treatment**

To assess the impact of the type of treatment (convalescent plasma/vaccine-boosted plasma/standard of care), a Cox regression model adjusting for type of treatment, and stratified for patient group is conducted. The hazard ratios and 95% profile-likelihood confidence intervals for “convalescent plasma vs standard of care”, “vaccine-boosted plasma vs standard of care” and “convalescent plasma vs vaccine-boosted plasma” will be obtained from the model. Patients who did receive both convalescent and vaccine-boosted plasma will be analysed together with those patients who received only vaccine-boosted plasma. Additionally, the cumulative improvement curves are calculated using the Kaplan-Meier method together with 95% log-log-type confidence bands, and will be calculated separately for the three types of treatment.

**7.5.3 Impact of type of COVID infection wave**

To assess the impact of the type of COVID infection wave (second wave: 3.9.2020 – 14.02.2021/third wave: 15.02.2021- 27.06.2021/fourth wave: since 28.06.2021), the cumulative improvement curves per treatment and patient group for each wave are calculated using the Kaplan-Meier method together with 95% log-log-type confidence bands.

**7.5.4 Impact of reception of monoclonal antibodies**

In case that more than 10% of all patients received monoclonal antibodies, the impact of this treatment will be assessed by calculating the cumulative improvement curves per treatment and patient group for patients with and without monoclonal antibody treatment using the Kaplan-Meier method together with 95% log-log-type confidence bands, and will be calculated separately for the three waves of treatment.

**7.5.5** **Impact of virus variant**

To assess the impact of the type of virus variant (Wild-type/Alpha/Delta/Omikron), the cumulative improvement curves per treatment and patient group for each variant are calculated using the Kaplan-Meier method together with 95% log-log-type confidence bands.

**7.5.6 Impact of vaccination status**

To assess the impact of the type of vaccination status (vaccinated/not vaccinated), the cumulative improvement curves per treatment and patient group for vaccinated and non-vaccinated patients are calculated using the Kaplan-Meier method together with 95% log-log-type confidence bands. Patients will be considered as vaccinated if they completed the cycle as per definition of the vaccine (1 vaccination for Johnson & Johnson vaccine, 2 vaccinations otherwise).

**7.5.7 Impact of number of neutralizing antibodies at baseline**

To assess the impact of the number of neutralizing antibodies at baseline (<1:40, ≥1:40), the cumulative improvement curves per treatment and patient group for each variant are calculated using the Kaplan-Meier method together with 95% log-log-type confidence bands.

Furthermore, the number of neutralizing antibodies at baseline will be included as continuous covariate in a Cox regression model additionally adjusting for treatment group, while stratifying for patient group. The hazard ratios and 95% profile-likelihood confidence intervals will be obtained from this model.

**7.6 Analysis of the secondary endpoints**

For all secondary endpoints, the analysis will be conducted based on the FAS and as a supplementary analysis based on the PPS.

**7.6.1 Analysis of overall survival**

Overall survival is analyzed by using a Cox regression model adjusted for the factor treatment and stratified by patient group, determining hazard ratios with 95% profile-likelihood confidence intervals and (descriptive) p-values. Survival estimates are calculated using the Kaplan-Meier method together with 95% log-log-type confidence bands, and the 28-day survival rate will be given for both treatment groups together with 95% log-log type confidence intervals. In addition, overall survival curves will also be calculated separately for each patient group per treatment group, and descriptive log-rank tests will be done to compare the two treatment groups for each patient group.

Analogously as for the supplementary analysis of the primary endpoint, an IPCW model and a RPSFT model will additionally be fitted for the overall survival to assess the potential treatment effect in the hypothetical scenario where patients did not switch from the control to the experimental arm.

**7.6.2 Analysis of time until discharge**

Time until discharge will be analyzed using a log-rank test stratified for the factor “patient group” and censoring patients who died from any cause at day 84 as described in chapter 7.3. Kaplan-Meier estimates for the cumulative discharge rate for both treatment groups together with 95% log-log-type confidence bands will be computed.

A Cox regression model, stratified for patient group, will be conducted to obtain an estimate for the hazard ratio for the treatment and a corresponding 95% profile-likelihood confidence interval.

IPCW and a RPSFT model will additionally be fitted for the endpoint ‘time until discharge’ to assess the potential treatment effect in the hypothetical scenario where patients did not switch from the control to the experimental arm.

**7.6.3 Analysis of requirement of mechanical ventilation**

The endpoint “requirement of mechanical ventilation (yes/no)” is analyzed by means of a logistic regression model adjusting for the factors treatment and patient group. If a patient who did not previously receive mechanical ventilation dies, this patient will be considered as having required mechanical ventilation. An estimate for the odds ratio for the factor treatment together with a 95% profile-likelihood confidence interval will be obtained from the model.

Also, absolute and relative frequencies will be given for this endpoint, together with 95% confidence intervals.

**7.6.4 Analysis of SARS-CoV-2 viral load and antibody titres**

Viral clearance (defined as having a viral load that is either non-measurable or <33) will be determined for every follow-up visit. Patients who died before experiencing a viral clearance will be regarded as non-cleared. Viral clearance at specific time points will be compared between the two treatment groups using a logistic regression model adjusting for the factors treatment and patient group.

Ct values for viral load will be determined as Ct=–log(viral load at follow-up/viral load at baseline) for days 5 and 10. Nonparametric van Elteren test will be used to compare the two treatment groups with regard to the Ct values while stratifying for patient group.

OD ratio and antibody titres will be evaluated on day 3 and 5 combined. In case that a measurement exists on both day 3 and day 5, the maximum will be used for analysis. The day 3/day 5 value will be descriptively assessed and compared to baseline. Nonparametric van Elteren test will be used to compare the two treatment groups with regard to the OD ratio and antibody titres while stratifying for patient group. Boxplots will be given for all time points for each treatment group to graphically investigate the Ct values, OD ratio, and antibody titres.

**7.6.5 Analysis of cytokine levels**

The cytokine levels will only be analysed descriptively. The ratio of the cytokine level at day 3/ day 4 / day 5 to the cytokine level at baseline will be calculated, and descriptive measures will be given for each treatment group.

**7.7 Analysis of exploratory endpoints**

All exploratory endpoints will be analyzed descriptively using the methods described in chapter 7.1. For endpoints that are assessed at multiple time points, each time point will be analyzed separately and line plots over time will be generated. The line plots will show the time point on the x-axis and either the mean value per group for parametrically evaluated continuous endpoints, the median value per group for all non-parametrically evaluated continuous and ordinal endpoints or the relative frequency per group for binary endpoints on the y-axis.

The exploratory endpoint SOFA score will be analysed following the recommendations of Lambden et al. (2019). The highest possible score of 24, corresponding to the worst outcome, will be used for patients who died before the SOFA score could be assessed during follow-up. In case that one or more items of a SOFA assessment are missing, mean imputation will be used to derive the total score. In case that all items are missing, no imputation will be done.

**7.8 Analysis of safety endpoints**

The assessment of safety is based mainly on the frequency of adverse events and on the laboratory values Safety analysis is based on the safety population. Adverse events are summarized by presenting the number and percentage of patients having any adverse events or serious adverse events, and having each individual adverse event, and by determining and summarizing the maximum individual toxicity grade (over all forms of toxicity). Furthermore, the most common AEs (those occurring in at least 10% of the treatment group) are determined. Event rates are summarized along with two-sided Clopper-Pearson 95% confidence intervals and analyzed by (descriptive) chi-squared tests. Any other information collected (e.g. severity or relatedness to study drug) are summarized using the descriptive methods described in chapter 7.1.

All laboratory values on day 5 will be descriptively evaluated as continuous measures and tabulated per treatment group. To compare laboratory values between treatment groups, two-sample t-tests will be conducted. In case a p-value smaller or equal to 0.05 is achieved, boxplots will additionally be given for this respective laboratory value.

**7.9 Subgroup analyses**

The primary endpoint and all secondary endpoints will be separately assessed for patient group 1, as well as for patient groups 1+2 combined.

**8 Differences to trial protocol**

Instead of recruiting the planned number of 174 patients, only 134 patients were enrolled to the trial as recruitment was stopped on January 20, 2022. The reason was a suspected inefficiency of the collected plasma bags for patients infected with the Omicron variant.

As viral load and antibody titres are not assumed to be normally distributed, they will be analysed using descriptive and non-parametric methods with no imputation being done for these endpoints.

**9 Interpretation of results**

As the planned number of n=174 was not reached, the study does not have the aspired power of 80% to detect the assumed difference between the two treatment groups with respect to the primary endpoint.

**10 Data problems**

In case that less than 60% of virus variants can be determined, a subgroup analysis based on this predictor (laid out in section 7.5.5) will not take place.

**11 Estimands framework**

In the Addendum to the ICH E9 guideline (final version), the estimands framework is recommended as clear and transparent definition of “what is to be estimated” (International Council for Harmonization 2019). The estimand is a construct of the treatment, the targeted population, the variable, a specification of how intercurrent events are handled, and a population-level summary measure.

**11.1 Main estimand**

The *main estimand* is described (referring to the primary efficacy analysis):

**Treatment:** Infusion of CP/PVP (on two days of intervention from two different donors) vs. standard of care

**Population:** The targeted population is defined through the in- and exclusion criteria.

**Variable:** Time from randomisation to clinical improvement within 84 days by two points on a seven point ordinal scale (as defined in chapter 5.1) or live discharge from the hospital

**Intercurrent events:**

- live discharge from the hospital is incorporated into the variable definition (composite strategy)
- death from any cause within 84 days after randomization without previous improvement is taken into account by censoring deceased patients at day 84 (composite strategy)
- treatment switch will be ignored (treatment policy strategy)

**Population-level summary:** Hazard ratio

**12 Software**

SAS Version 9.4 will be used for all statistical analyses. R (<http://r-project.org>) version 4.0.0 or higher as well as ggplot2 (Wickham 2016) can be used for the creation of figures.

2. **Figures**

Figure S1 Consort diagram for group 1 (hematological and solid cancer)

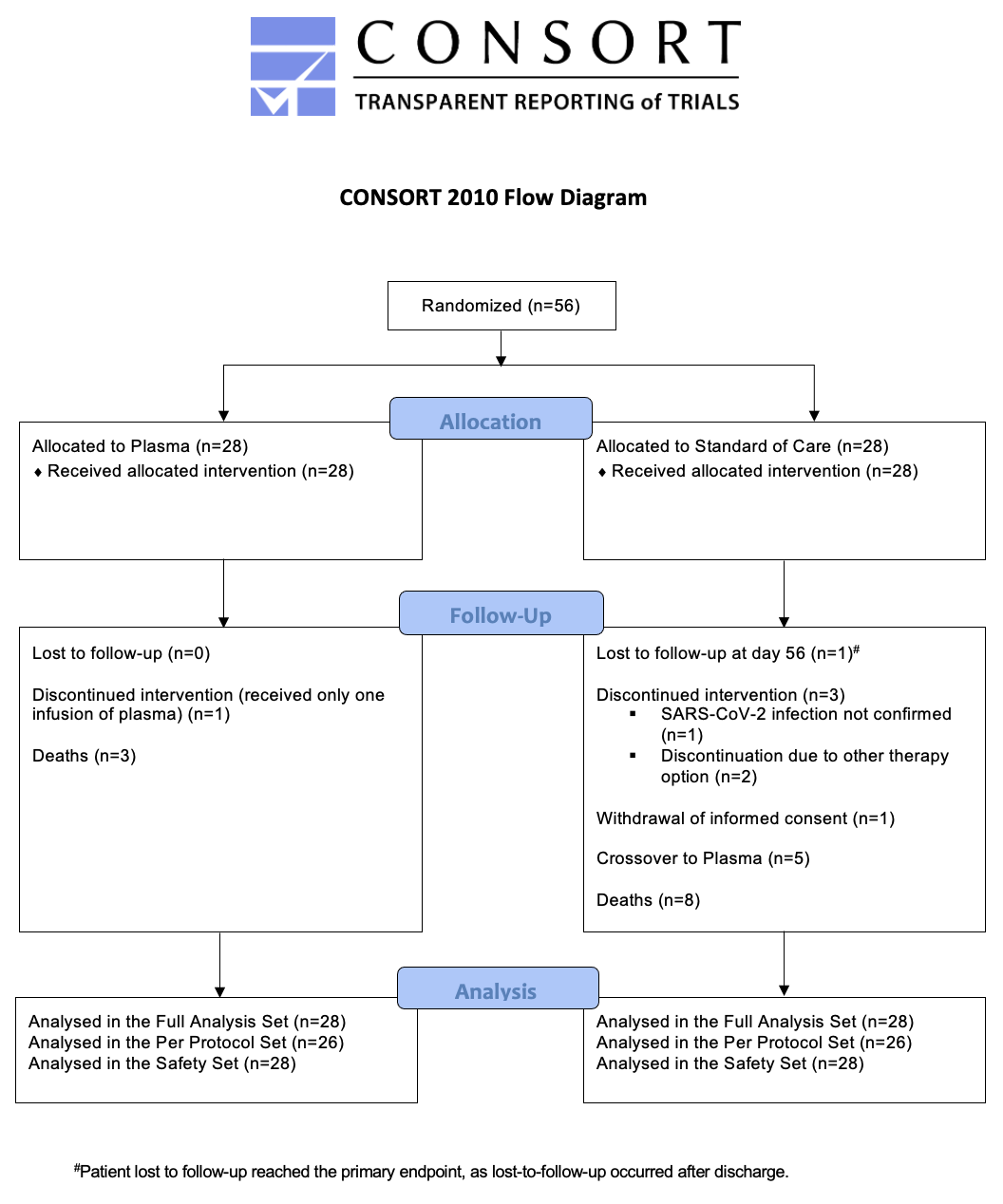

Figure S2 Kaplan Meier curves for time to improvement on 7-point ordinal scale or live hospital discharge for groups-2 to -4

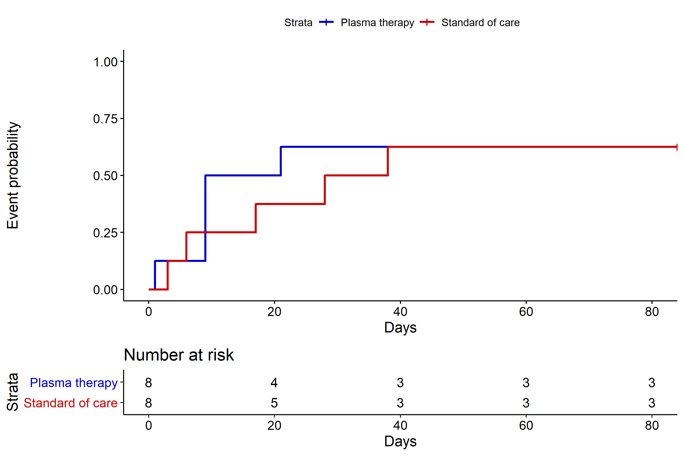

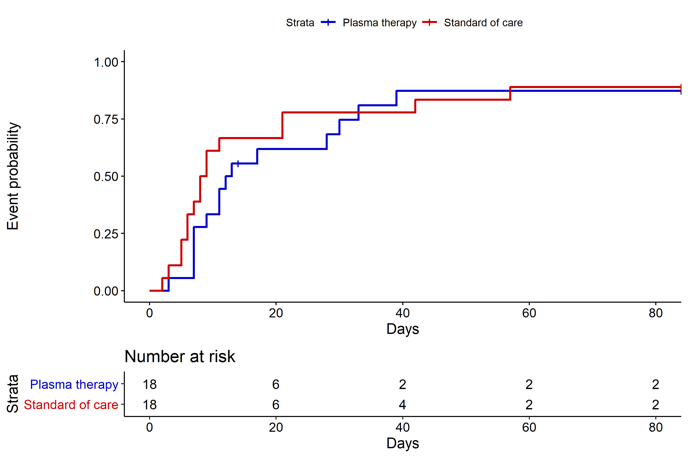

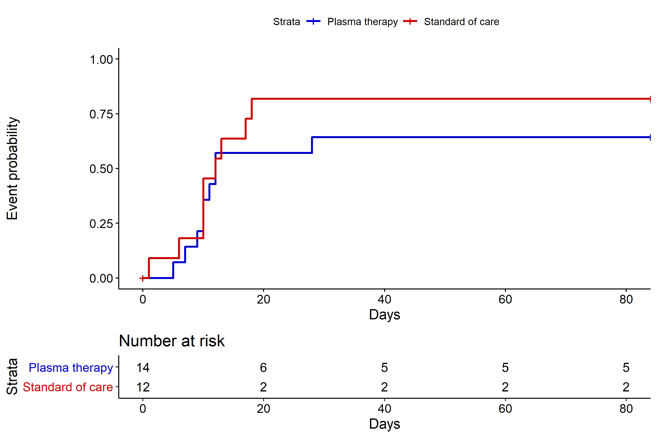

Kaplan Meier curves for cumulative probability of endpoint 2-point improvement or discharge on the 7-point ordinal scale with number of subjects at risk below

Left: group 2; center: group 3; right: group 4

Figure S3 Kaplan Meier curves for Survival for groups 2-4

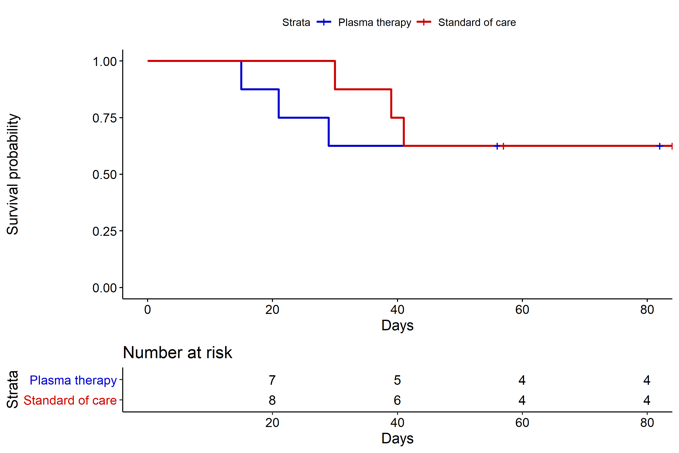

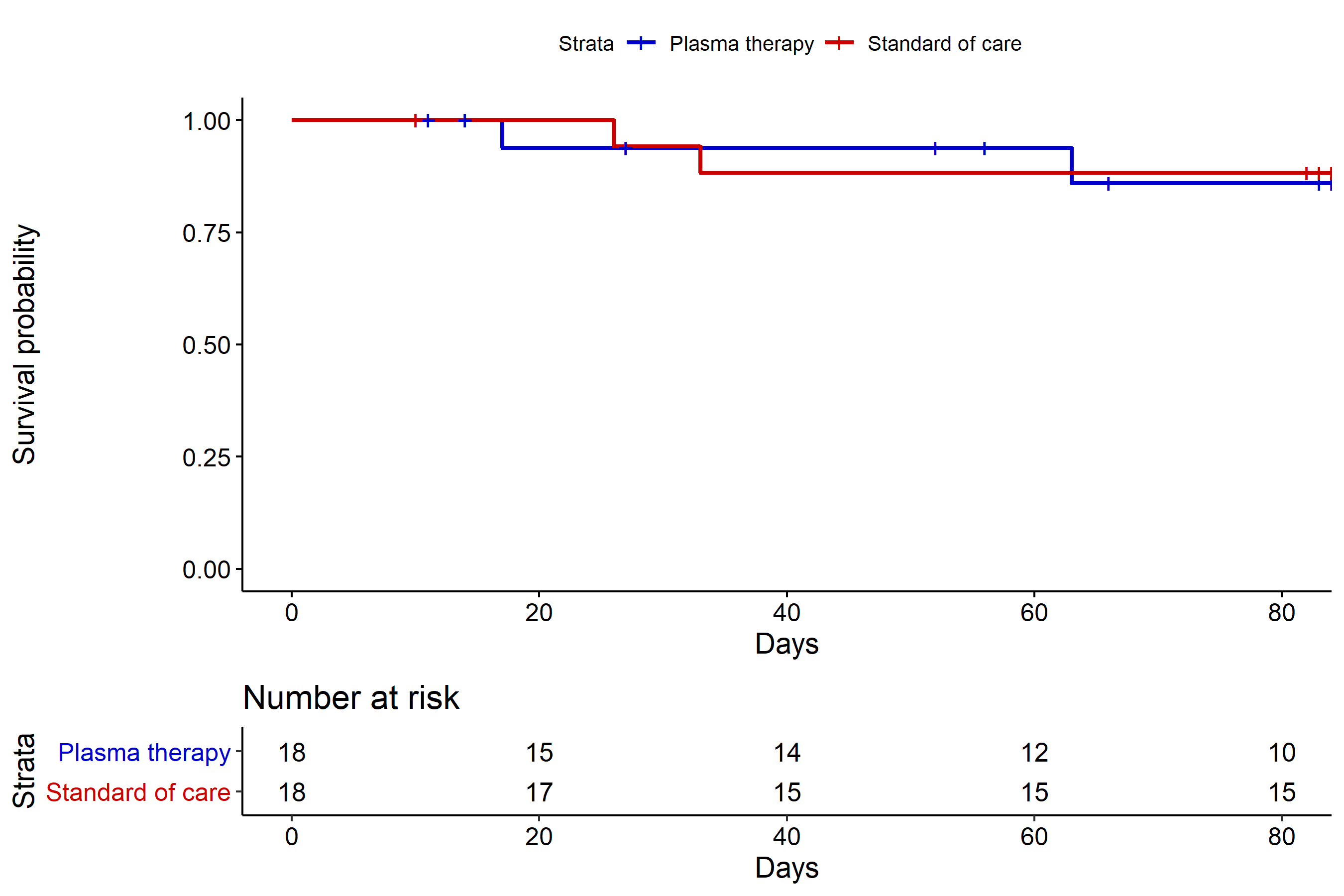

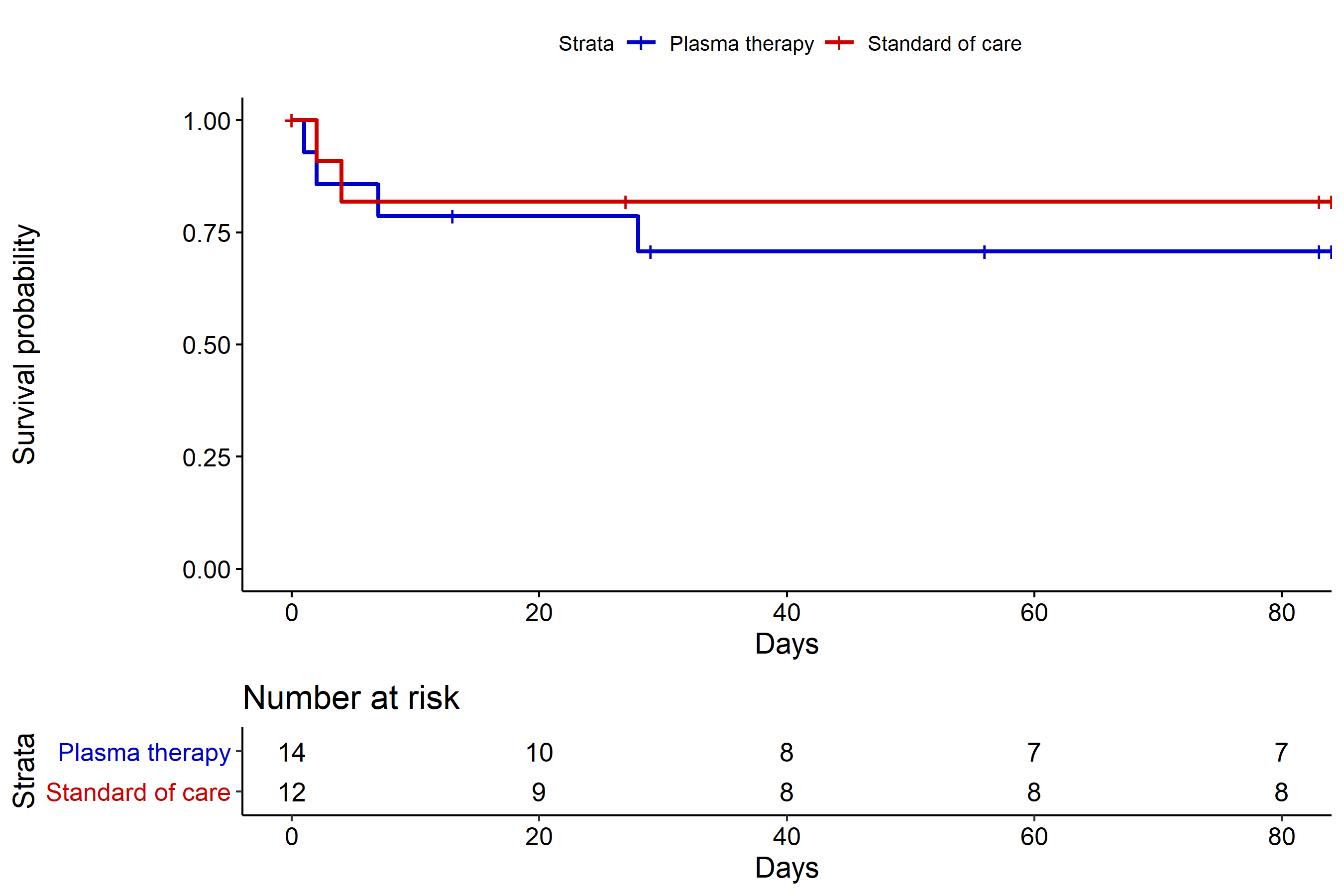
Kaplan Meier curves for endpoint event survival with number of subjects at risk

Left: group 2; center: group 3; right: group 4

Figure S4 Kaplan Meier curves for ‘Time to discharge’ overall

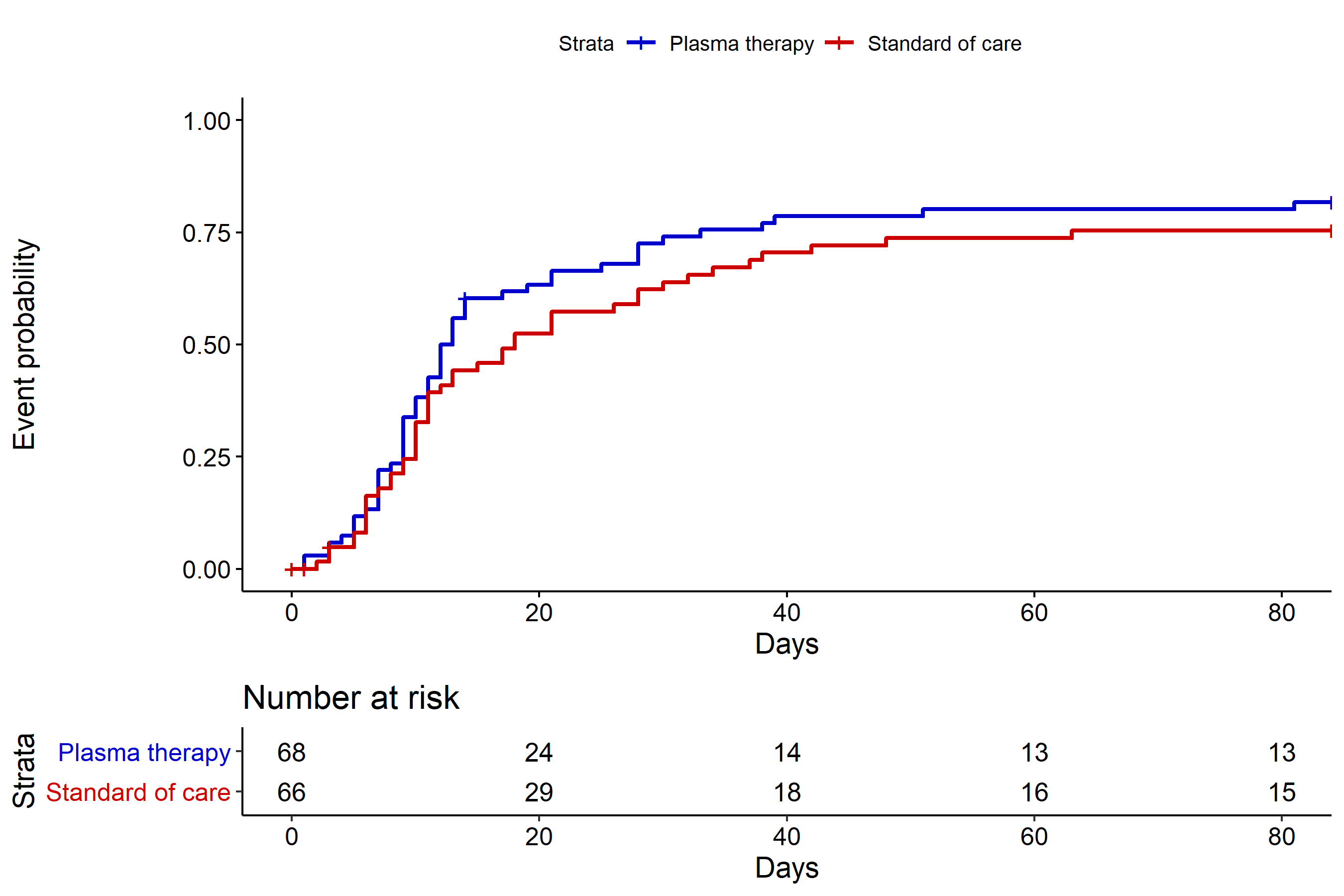

Kaplan Meier curves for cumulative endpoint event discharge from hospital with number of subjects at risk (above x-axis)

Figure S5: NeutraLISA measurement in the four patient groups

(A) Measurement at baseline (day 1)

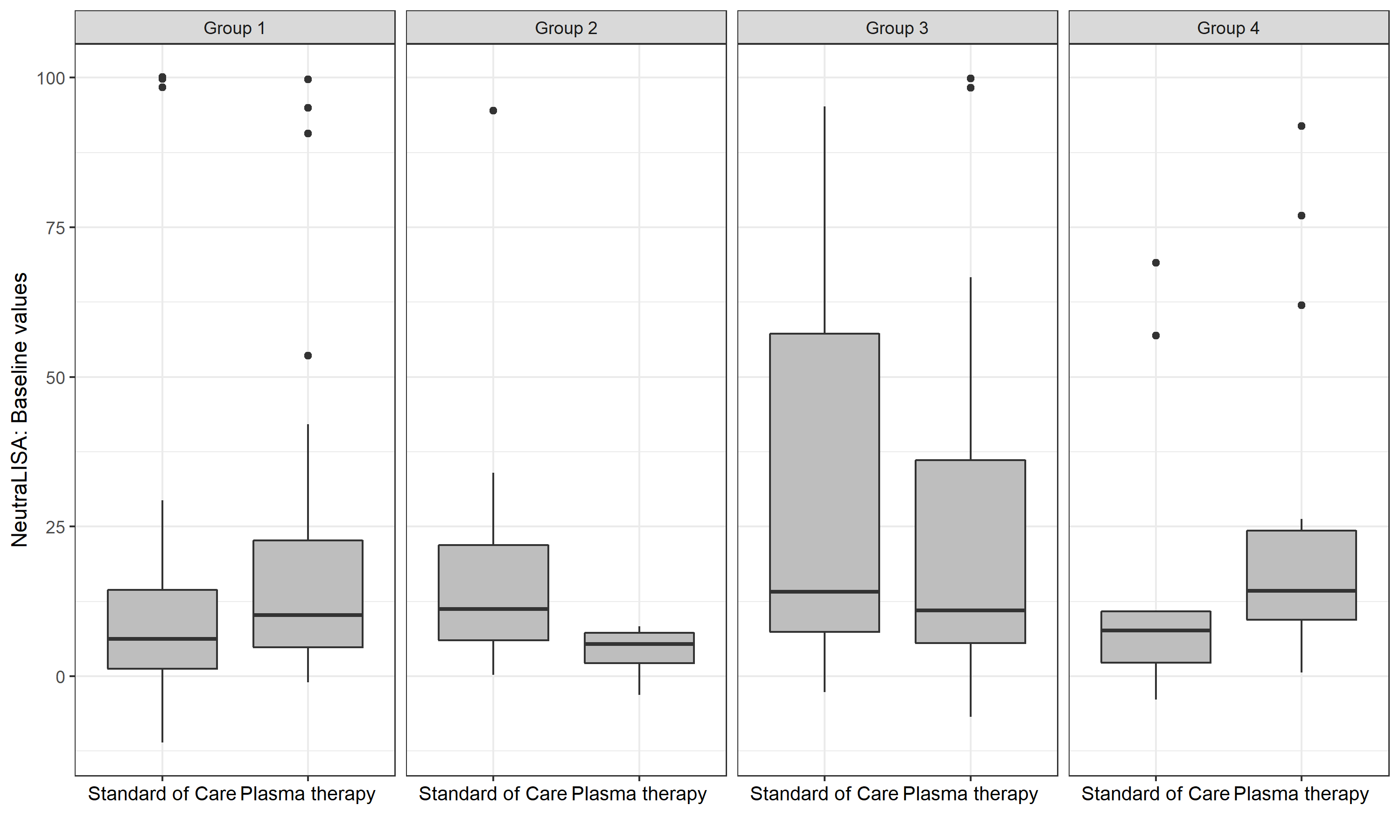

(B) Maximum measurement at day 3/5

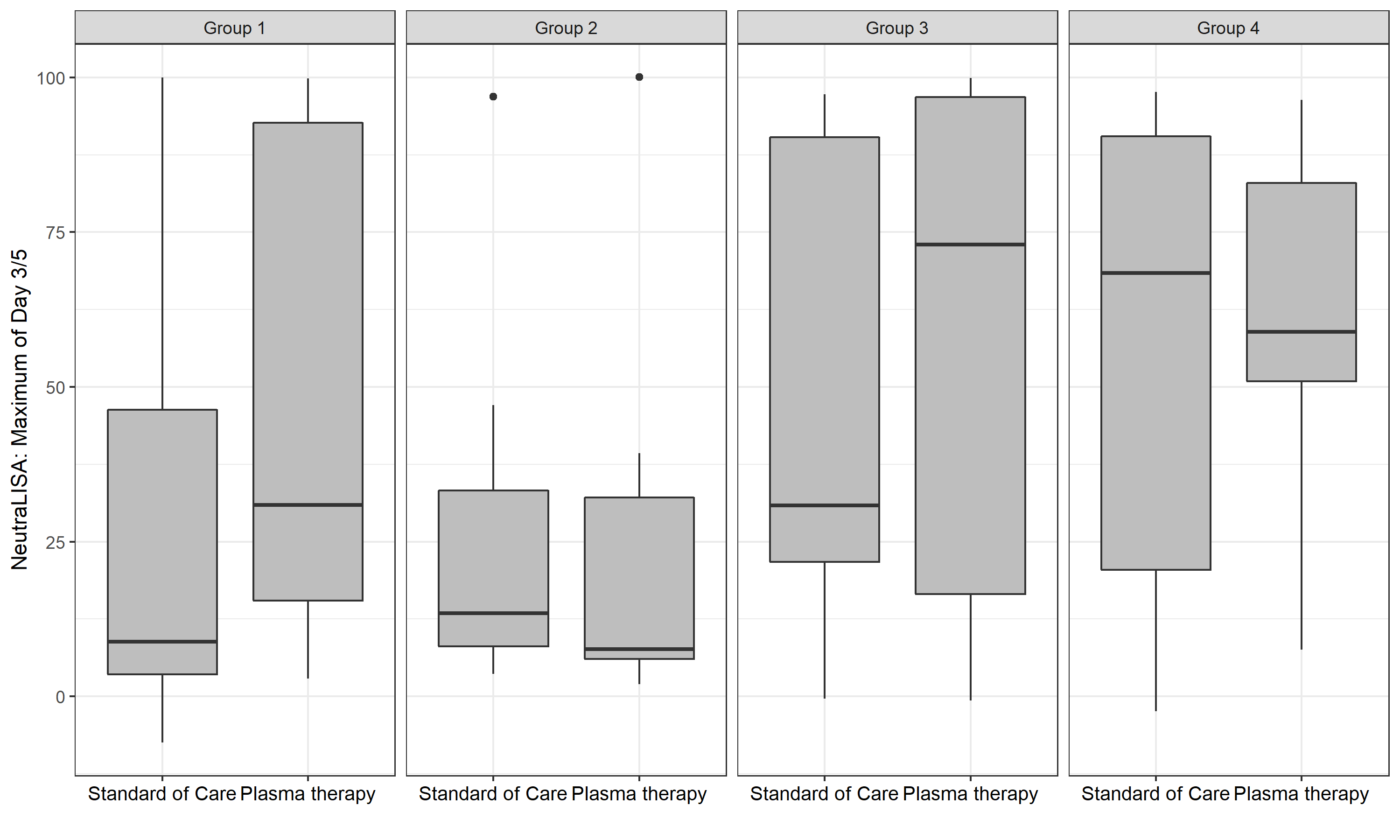

(C) Difference between Day 1 and Day 3/5

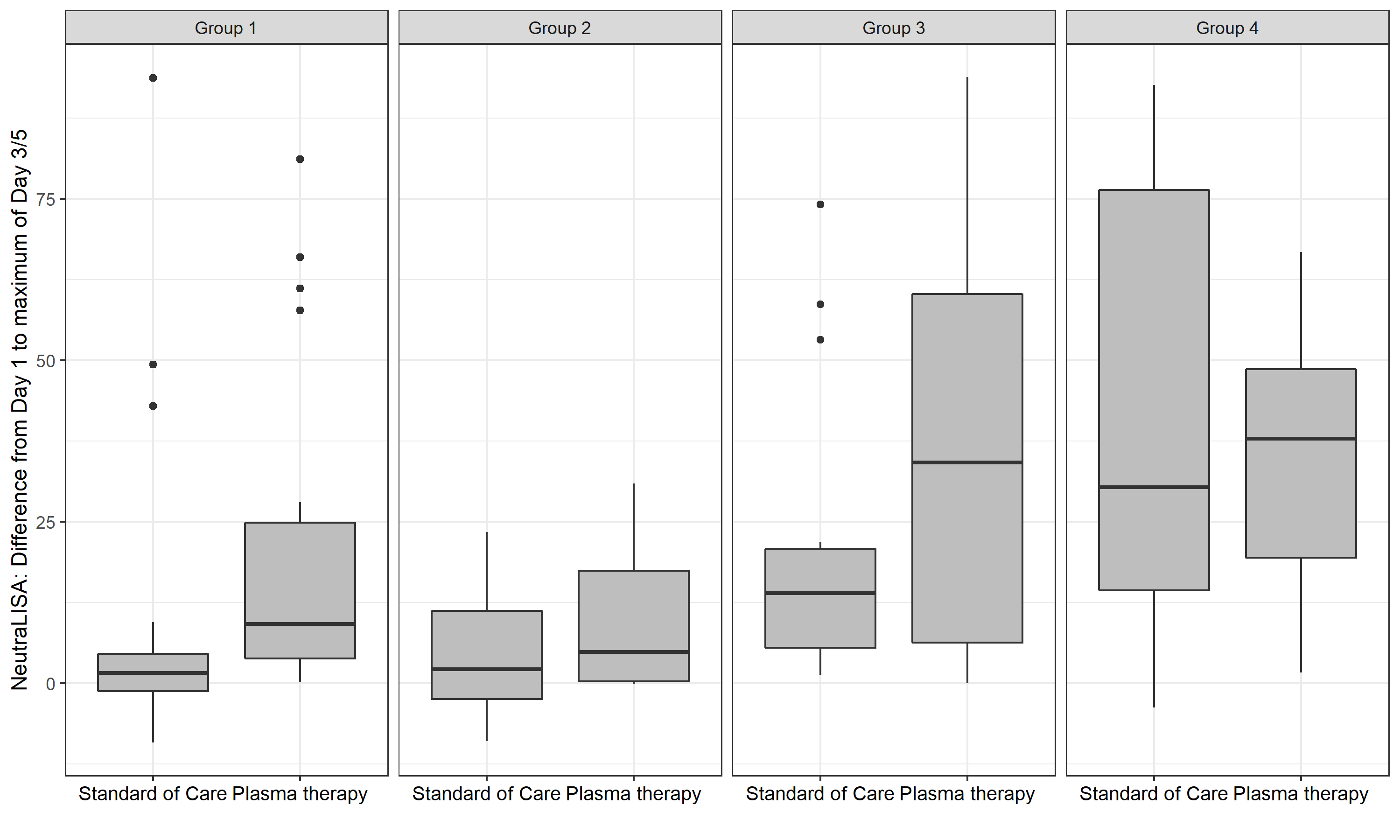

Figure S6: Column scatter plots for neutralization characteristics of plasma donors

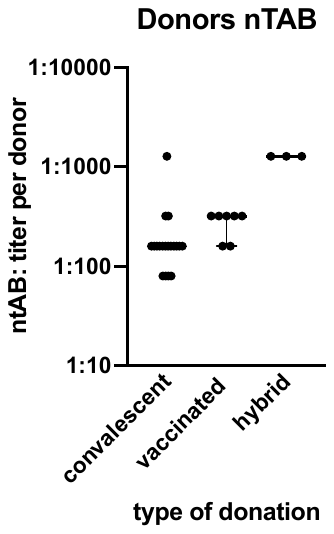

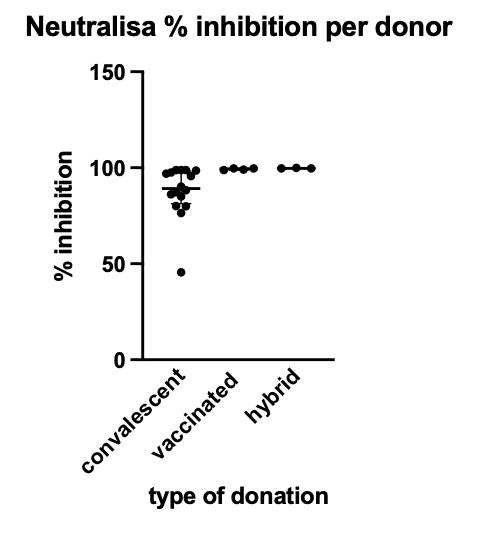

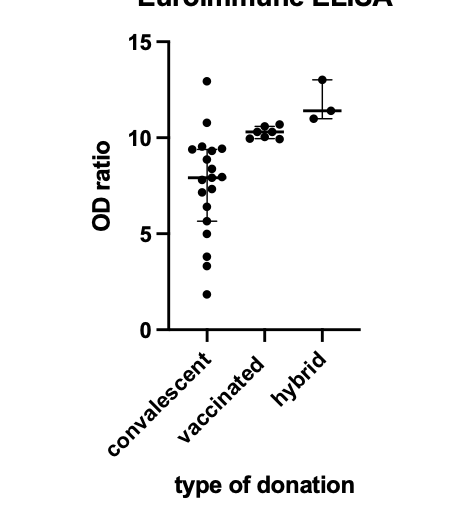

Upper left: ntAB titers per plasma donor; upper right: neutraLISA assay result as % inhibition

Abbreviations: ntAB neutralizing Antibodies; lower left: Euroimmune ELISA values

Figure S7: Correlation between live virus neutralization assay and NeutraLISA in donor plasma

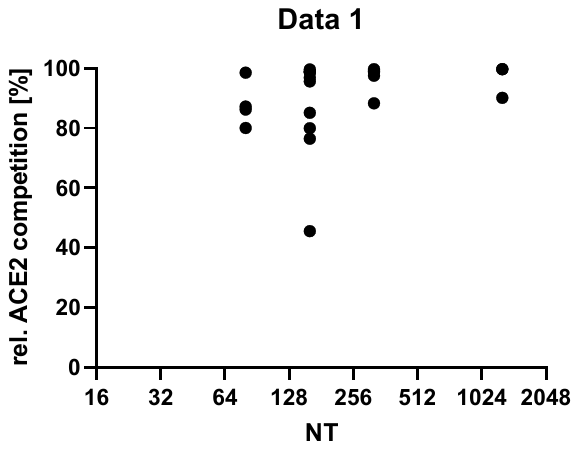

Caption: NeutraLISA results on y-axis and live virus neutralization assay (as described in text S3) on the x-axis. Results show saturation of ACE2 competition in the NeutraLISA with titers of 1:80 and above.

3. **Tables**

Table S1 Trial sites, enrolled patients and local investigators

| Trial sites | Principal Investigators | Enrolled patients, n=134 |
| --- | --- | --- |
| Department of Internal Medicine V, Heidelberg University Hospital, Heidelberg | Carsten Müller-Tidow  Uta Merle  Johann Motsch | 45 |
| Department of Internal Medicine III, Klinikum Chemnitz gGmbH, Chemnitz | Mathias Hänel  Anke Morgner | 13 |
| Medical Department I, Klinikum Bremen-Mitte, Bremen | Bernd Hertenstein  Rolf Dembinski | 9 |
| Department of Medicine II, Division of Infectious Diseases and Travel Medicine, University Medical Centre Freiburg, Freiburg | Winfried V. Kern  Dirk Wagner | 9 |
| Department of Internal Medicine, Infectious Diseases, University Hospital Frankfurt, Goethe University Frankfurt, Frankfurt am Main | Timo Wolf  Maria J.G.T. Vehreschild | 8 |
| Department of Hematology, Oncology, Palliative Care and Stem Cell Transplantation, Klinikum Hochsauerland GmbH, Meschede | Mohammad-Amen Wattad  Elisabeth Lange | 8 |
| I. Department of Medicine, University Medical Center Hamburg-Eppendorf, Hamburg | Stefan Schmiedel  Marylyn M. Addo | 7 |
| Department of Hematology, Oncology and Tumor Immunology, Charité University Medicine, Campus Virchow Clinic, Berlin | Lars Bullinger  Jörg Westermann | 6 |
| Department of Internal Medicine II, Klinikum Darmstadt GmbH, Darmstadt | Carl C. Schimanski  Jens Kittner  Frank Staib | 6 |
| Pneumology and Critical Care Medicine, Thoraxklinik, University of Heidelberg, Heidelberg | Felix Herth  Michael Kreuter | 6 |
| Department of Internal Medicine I, University Hospital and Faculty of Medicine Carl Gustav Carus of TU Dresden, Dresden | Nael Alakel  Martin Bornhäuser | 4 |
| Department of Infectious Diseases, West German Centre of Infectious Diseases, University Hospital Essen, University Duisburg-Essen | Oliver Witzke  Sebastian Dolff | 4 |
| Department I of Internal Medicine, Frankfurt (Oder) General Hospital, Frankfurt/Oder | Olaf Hopfer  Michael Kiehl | 4 |
| Department of Medicine B, Gastroenterology and Hepatology, University Hospital Münster, Münster | Phil-Robin Tepasse  Hartmut Schmidt  Viorelia Stoica | 3 |
| Department of Internal Medicine I, Klinikum Herford | Matthias Ruhe  Jan Kähler | 2 |
| TOTAL |  | N=134 |

Table S2 Protocol amendments and SAP deviations

|  | *Content* | *Justification* | *Date of change* |
| --- | --- | --- | --- |
| *Changes to the protocol* | In-/Exclusion Criteria:  Blood hemoglobin concentration changed from ≥ 10 g/dl to ≥ 8 g/dl.  (Change protocol V1.2 🡪 V2.0) | Main concern for limitation was the amount of blood collections from the trial. With decreasing blood collections, we also decreased the hemoglobin cut-off to increase the number of eligible patients, particularly those with preexisting hematological malignancies | 10^th^ November 2020 |
|  | Trial Schedule:  The Trial Schedule was revised by reducing the number of assessments, blood collections and follow-up tests.  (Change protocol V1.2 🡪 V2.0) | to ensure a smooth conduct of the study by the study teams and to expose patients to less follow-up treatments. The revision was done by ensuring patient´s safety without impacting the study endpoints. | 10^th^ November 2020 |
|  | Plasma product:  1. Addition of the use of plasma of vaccinated donors (Updated IMP name: RECOVER CT SARS-CoV-2-ab Plasma COVID-19 (HD): Patients can receive plasma either from convalescents and/or from vaccinated persons. (Change protocol V2.0 🡪 V3.0) | With data becoming available of increased titers of antibodies in vaccinated donors and increased numbers of people being vaccinated (i.e. increased access to donors), we considered vaccinated donors in addition to convalescent donors | 30^st^ March 2021 |
|  | Number of Trial Sites: expanded to 10-15 medical centers. (Change protocol V1.2 🡪 V2.0) | to increase enrollment and speed of enrollment | 10^th^ November 2020 |
| *Changes to the SAP* | Requirement of mechanical ventilation:  Patients with a follow-up time of at most one day were excluded from the analysis, this was not pre-specified. | Duration was considered too short for outcome to occur | At time of analysis |

Table S3 Representativeness

| **Category** |  |
| --- | --- |
| Disease | Severe Covid-19 in patients with underlying disease and risk factors |
| Sex and gender | COVID-19 affects men and women similarly. Severe COVID-19 is more frequent in men than in women. |
| Age | Severe COVID-19 as well as the underlying risk groups 1 and 2 defined in the trial increase in frequency with age. |
| Ethnic group | Ethnic groups were represented within the RECOVER trial as indicated in Supplementary Table 1. |
| Geography | No differences within Germany were considered. Patients were included in University Hospitals and in larger community hospitals to ensure adequate trial representation. |
| Other considerations | In many clinical trials, patients above 75 years of age are often excluded. The RECOVER trial specifically set up group 4 to allow study participation of this vulnerable group of COVID-19 patients. |
| Overall representativeness of this trial | Men and women were recruited into this trial with about 1/3 of trial participants being women. Due to specific considerations for patients with advanced age, the median age was higher compared to other clinical trials for severe COVID-19. |

Table S4 Plasma donor characteristics

|  | Convalescent  donor (CP) | Vaccinated donor (VP) | Convalescent and vaccinated donor (C+VP) | all |
| --- | --- | --- | --- | --- |
| n | 19 | 7 | 3 | 29 |
| Gender, female/male (%) | 6/13  20.69/44.83 | 5/2  17.24/6.90 | 0/3  0/10.34 | 11/18  37.93/62.07 |
| Age [years] - median (IQR) | 46 (31-52) | 26 (25-39) | 31 (29-56) | 39 (32-43) |
| Euroimmune ELISA OD – median (IQR) | 7.92 (5.65-9.39) | 10.30 (9.96-10.60) | 11.40 (10.99-13.02) | 10.26 (7.52-11.80) |
| Neutralization titer median (IQR) | 1:160 (1:160-1:160) | 1:320 (1:160-1:320) | 1:1280 (1:1280-1:1280) | 1:274 (1:219-1:1280) |
| Neutralization determined by NEUTRALISA surrogate assay [%]  median (IQR) | 89.23 (81.32-98.33)^+^ | 99.38 (98.85-99.68)^++^ | 99.69 (99.69-99.91) | 99.30 (87.76-99.76) |
| Missing: ^+^n = 3, ^++^n=3  Abbreviations:  CP=convalescent plasma, no vaccination  VP=vaccinated plasma, 2 doses of mRNA vaccine against SARS-CoV-2, no known infection with SARS-CoV-2  C+VP=convalescent plasma and at least 1 mRNA vaccine boost against SARS-CoV-2 | | | | |

Table S5 Epidemiological data of study participants in predefined subgroups

|  | **Group 1: Hematological and solid Cancer** | | |
| --- | --- | --- | --- |
|  | all (n=56) | Control (n=28) | Plasma (n=28) |
| General | | | |
| Age mean +/- SD | 66.7 +/-10.5 | 70.6 +/-8.4 | 62.9 +/-11.1 |
| Gender |  |  |  |
| male | 37 (66.1%) | 16 (57.1%) | 21 (75.0%) |
| female | 19 (33.9%) | 12 (42.9%) | 7 (25.0%) |
| Ethnic origin |  |  |  |
| Asian | 0 (0.0%) | 0 (0.0%) | 0 (0.0%) |
| Caucasian/White | 54 (96.4%) | 28 (100.0%) | 26 (92.9%) |
| Hispanic | 2 (3.6%) | 0 (0.0%) | 2 (7.1%) |
| Other | 0 (0.0%) | 0 (0.0%) | 0 (0.0%) |
| Time from symptom onset to randomization (days) (median (p25,p75)) | 6.5 (4.0, 11.5) | 6.0 (4.0, 10.0) | 7.0 (4.0, 12.0) |
| Comorbidities |  |  |  |
| chronic lung disease | 11 (19.6%) | 4 (14.3%) | 7 (25.0%) |
| cardiovascular disease | 35 (62.5%) | 20 (71.4%) | 15 (53.6%) |
| chronic liver disease | 5 (8.9%) | 5 (17.9%) | 0 (0.0%) |
| Rheumatic / immunologic disease | 5 (8.9%) | 3 (10.7%) | 2 (7.1%) |
| Organ transplant | 5 (8.9%) | 3 (10.7%) | 2 (7.1%) |
| Diabetes | 18 (32,1%) | 12 (42,9%) | 6 (21,4%) |
| Chronic kidney disease | 12 (21.4%) | 7 (25.0%) | 5 (17.9%) |
| *with hemodialysis* | 4 (7.1%) | 3 (10.7%) | 1 (3.6%) |
| Clinical Frailty Scale (median (p25,p75))* | 3.0 (2.0, 4.0) | 3.0 (3.0, 4.0) | 4.0 (2.0, 4.0) |
| WHO Performance Status* |  |  |  |
| ECOG =0 | 3 (5.5%) | 1 (3.7%) | 2 (7.1%) |
| ECOG =1 | 12 (21.8%) | 5 (18.5%) | 7 (25.0%) |
| ECOG =2 | 25 (45.5%) | 13 (48.1%) | 12 (42.9%) |
| ECOG =3 | 11 (20.0%) | 5 (18.5%) | 6 (21.4%) |
| ECOG =4 | 4 (7.3%) | 3 (11.1%) | 1 (3.6%) |
| Cancer (pre-existing or concurrent hematological malignancy and/or active cancer therapy (incl. chemotherapy, radiotherapy, surgery) within the last 24 months or less) | | | |
| all entities | 56 (41,8%) | 28 (42,4%) | 28 (41,2%) |
| B-NHL/CLL | 18 (32.1%) | 7 (25.0%) | 11 (39.3%) |
| AML/MDS | 12 (21.4%) | 8 (28.6%) | 4 (14.3%) |
| Myeloma | 11 (19.6%) | 6 (21.4%) | 5 (17.9%) |
| B-ALL | 2 (3.6%) | 0 (0.0%) | 2 (7.1%) |
| Hodgkin | 2 (3.6%) | 1 (3.6%) | 1 (3.6%) |
| CML | 1 (1.8%) | 1 (3.6%) | 0 (0.0%) |
| T-NHL | 1 (1.8%) | 1 (3.6%) | 0 (0.0%) |
| solid tumor | 9 (16.1%) | 4 (14.3%) | 5 (17.9%) |

*n≠56

Table S5 Epidemiological data of study participants in predefined subgroups (continued)

|  | **Group 2: Immunosuppression** | | |
| --- | --- | --- | --- |
|  | all (n=16) | Control (n=8) | Plasma (n=8) |
| General | | | |
| Age mean +/- SD | 61.2 +/-13.6 | 58.0 +/-9.8 | 64.4 +/-16.6 |
| Gender |  |  |  |
| male | 14 (87.5%) | 7 (87.5%) | 7 (87.5%) |
| female | 2 (12.5%) | 1 (12.5%) | 1 (12.5%) |
| Ethnic origin |  |  |  |
| Asian | 0 (0.0%) | 0 (0.0%) | 0 (0.0%) |
| Caucasian/White | 15 (93.8%) | 7 (87.5%) | 8 (100.0%) |
| Hispanic | 0 (0.0%) | 0 (0.0%) | 0 (0.0%) |
| Other | 1 (6.3%) | 1 (12.5%) | 0 (0.0%) |
| Time from symptom onset to randomization (days) (median (p25,p75)) | 9.5 (6.0, 12.5) | 12.5 (10.0, 17.0) | 6.5 (3.0, 9.0) |
| Comorbidities |  |  |  |
| chronic lung disease | 5 (31.3%) | 2 (25.0%) | 3 (37.5%) |
| cardiovascular disease | 13 (81.3%) | 6 (75.0%) | 7 (87.5%) |
| chronic liver disease | 4 (25.0%) | 1 (12.5%) | 3 (37.5%) |
| Rheumatic / immunologic disease | 7 (43.8%) | 3 (37.5%) | 4 (50.0%) |
| Organ transplant | 12 (75.0%) | 7 (87.5%) | 5 (62.5%) |
| Diabetes | 4 (25.0%) | 0 (0.0 %) | 4 (25.0%) |
| Chronic kidney disease | 10 (62.5%) | 5 (62.5%) | 5 (62.5%) |
| *with hemodialysis* | 0 (0.0 %) | 0 (0.0 %) | 0 (0.0 %) |
| Clinical Frailty Scale (median (p25,p75)) | 3.0 (2.5, 4.0) | 3.0 (2.5, 4.0) | 3.0 (2.5, 3.5) |
| WHO Performance Status |  |  |  |
| ECOG =0 | 0 (0.0 %) | 0 (0.0 %) | 0 (0.0 %) |
| ECOG =1 | 3 ( 18.8%) | 1 ( 12.5%) | 2 ( 25.0%) |
| ECOG =2 | 7 ( 43.8%) | 3 ( 37.5%) | 4 ( 50.0%) |
| ECOG =3 | 4 ( 25.0%) | 2 ( 25.0%) | 2 ( 25.0%) |
| ECOG =4 | 2 ( 12.5%) | 2 ( 25.0%) | 0 ( 0.0%) |

*n≠16

Table S5 Epidemiological data of study participants in predefined subgroups (continued)

|  | **Group 3: Lymphopenia/high D-dimers: aged >50 and ≤75 years** | | |
| --- | --- | --- | --- |
|  | all (n=36) | Control (n=18) | Plasma (n=18) |
| General | | | |
| Age mean +/- SD | 64.9 +/-7.2 | 64.9 +/-7.3 | 64.9 +/-7.3 |
| Gender |  |  |  |
| male | 27 (75.0%) | 15 (83.3%) | 12 (66.7%) |
| female | 9 (25.0%) | 3 (16.7%) | 6 (33.3%) |
| Ethnic origin |  |  |  |
| Asian | 1 (2.8%) | 1 (5.6%) | 0 (0.0%) |
| Caucasian/White | 35 (97.2%) | 17 (94.4%) | 18 (100.0%) |
| Hispanic | 0 (0.0%) | 0 (0.0%) | 0 (0.0%) |
| Other | 0 (0.0%) | 0 (0.0%) | 0 (0.0%) |
| Time from symptom onset to randomization (days) (median (p25,p75)) | 7.5 (6.0, 9.5) | 9.0 (4.0, 10.0) | 7.0 (6.0, 8.0) |
| Comorbidities |  |  |  |
| chronic lung disease | 9 (25.0%) | 5 (27.8%) | 4 (22.2%) |
| cardiovascular disease | 24 (66.7%) | 10 (55.6%) | 14 (77.8%) |
| chronic liver disease | 5 (13.9%) | 4 (22.2%) | 1 (5.6%) |
| Rheumatic / immunologic disease | 4 (11.1%) | 2 (11.1%) | 2 (11.1%) |
| Organ transplant | 0 (0.0 %) | 0 (0.0 %) | 0 (0.0 %) |
| Diabetes | 4 (11.1 %) | 1 (5.6 %) | 3 (16.7%) |
| Chronic kidney disease | 8 (22.2%) | 5 (27.8%) | 3 (16.7%) |
| *with hemodialysis* | 8 (22.2%) | 5 (27.8%) | 3 (16.7%) |
| Clinical Frailty Scale (median (p25,p75))* | 2.0 (1.0, 4.0) | 4.0 (2.0, 4.0) | 2.0 (1.0, 3.5) |
| WHO Performance Status |  |  |  |
| ECOG =0 | 0 (0.0 %) | 0 (0.0 %) | 0 (0.0 %) |
| ECOG =1 | 10 ( 27.8%) | 6 ( 33.3%) | 4 ( 22.2%) |
| ECOG =2 | 9 ( 25.0%) | 5 ( 27.8%) | 4 ( 22.2%) |
| ECOG =3 | 12 ( 33.3%) | 5 ( 27.8%) | 7 ( 38.9%) |
| ECOG =4 | 5 ( 13.9%) | 2 ( 11.1%) | 3 ( 16.7%) |

*n≠36

Table S5 Epidemiological data of study participants in predefined subgroups (continued)

|  | **Group 4: age >75 years** | | |
| --- | --- | --- | --- |
|  | all (n=26) | Control (n=12) | Plasma (n=14) |
| General | | | |
| Age mean +/- SD | 81.7 +/-4.3 | 82.3 +/-4.8 | 81.1 +/-4.0 |
| Gender |  |  |  |
| male | 13 (50.0%) | 8 (66.7%) | 5 (35.7%) |
| female | 13 (50.0%) | 4 (33.3%) | 9 (64.3%) |
| Ethnic origin |  |  |  |
| Asian | 0 (0.0%) | 0 (0.0%) | 0 (0.0%) |
| Caucasian/White | 26 (100.0%) | 12 (100.0%) | 14 (100.0%) |
| Hispanic | 0 (0.0%) | 0 (0.0%) | 0 (0.0%) |
| Other | 0 (0.0%) | 0 (0.0%) | 0 (0.0%) |
| Time from symptom onset to randomization (days) (median (p25,p75)) | 6.5 ( 3.0, 9.0) | 5.0 (2.0, 9.0) | 7.0 (6.0, 9.0) |
| Comorbidities |  |  |  |
| chronic lung disease | 12 (46.2%) | 9 (75.0%) | 3 (21.4%) |
| cardiovascular disease | 22 (84.6%) | 11 (91.7%) | 11 (78.6%) |
| chronic liver disease | 1 (3.8%) | 1 (8.3%) | 1 (3.8%) |
| Rheumatic / immunologic disease | 0 (0.0 %) | 0 (0.0 %) | 0 (0.0 %) |
| Organ transplant | 0 (0.0 %) | 0 (0.0 %) | 0 (0.0 %) |
| Diabetes | 8 (30,8%) | 6 (50 %) | 2 (14.3%) |
| Chronic kidney disease | 5 (19.2%) | 4 (33.3%) | 1 (7.1%) |
| *with hemodialysis* | 1 (3.8%) | 1 (8.3%) | 0 (0.0%) |
| Clinical Frailty Scale (median (p25,p75))* | 3.0 (3.0, 5.0) | 4.0 (2.5, 6.0) | 3.0 (3.0, 4.0) |
| WHO Performance Status* |  |  |  |
| ECOG =0 | 0 (0.0 %) | 0 (0.0 %) | 0 (0.0 %) |
| ECOG =1 | 2 (8.0%) | 0 (0.0%) | 2 (15.4%) |
| ECOG =2 | 10 (40.0%) | 8 (66.7%) | 2 (15.4%) |
| ECOG =3 | 8 (32.0%) | 4 (33.3%) | 4 (30.8%) |
| ECOG =4 | 5 (20.0%) | 0 (0.0%) | 5 (38.5%) |

*n≠26

Table S6 Baseline values and treatment in predefined subgroups

|  | **Group 1: Hematological and solid Cancer** | | |
| --- | --- | --- | --- |
|  | all (n=56) | Control (n=28) | Plasma (n=28) |
| SARS-CoV-2 Baseline | | | |
| Percent inhibition NeutraLISA (median (p25,p75))* | 8.1 (3.4, 17.3) | 6.3 (1.3, 15.2) | 10.2 (4.9, 22.7) |
| CT value on Day of randomization/Day 1 (Mean +/- SD)* | 22.2 +/-6.2 | 21.7 +/-5.9 | 22.7 +/-6.5 |
| Study assessments | | | |
| Seven-point ordinal scale at randomization |  |  |  |
| SPOS =3 | 12 (21.4%) | 5 (17.9%) | 7 (25.0%) |
| SPOS =4 | 38 (67.9%) | 20 (71.4%) | 18 (64.3%) |
| SPOS =5 | 6 (10.7%) | 3 (10.7%) | 3 (10.7%) |
| Laboratory (median (p25,p75)) |  |  |  |
| WBC (Gpt/l) | 4.7 (1.9, 7.3) | 4.4 (2.1, 8.1) | 4.8 (1.7, 6.4) |
| Lymphocytes (Gpt/l)* | 0.6 (0.2, 1.1) | 0.6 (0.2, 1.0) | 0.6 (0.3, 1.1) |
| CRP (mg/l)* | 91.2 (42.0, 171.0) | 84.9 (50.1, 171.0) | 93.2 (39.3, 166.1) |
| LDH (U/l)* | 323.5 (216.6, 457.0) | 331.1 (229.2, 473.1) | 301.0 (206.4, 457.0) |
| D-Dimer (mg/l)* | 1.4 (0.8, 2.4) | 2.2 (1.3, 3.5) | 1.2 (0.7, 2.1) |
| Troponin (pg/ml)* | 16.0 (11.0, 29.4) | 20.9 (13.8, 37.6) | 13.9 (10.5, 24.3) |
| Treatment (incl. cross-over day 10) | | | |
| Plasma received |  |  |  |
| Convalescent Plasma | 22 | 1 | 21 |
| Convalescent + Vaccinated plasma | 7 | 3 | 4 |
| Vaccinated plasma only | 4 | 1 | 3 |
| Other COVID-19 medication | | | |
| antiinflammatory | 19 (33.9%) | 8 (28.6%) | 11 (39.3%) |
| small-molecule antiviral | 4 (7.1%) | 0 (0.0%) | 4 (14.3%) |
| biologic antiviral | 3 (5.4%) | 1 (3.6%) | 2 (7.1%) |
| antibiotics | 0 (0.0%) | 0 (0.0%) | 0 (0.0%) |
| anticoagulants | 0 (0.0%) | 0 (0.0%) | 0 (0.0%) |
| other concomitant medication | 2 (3.6%) | 1 (3.6%) | 1 (3.6%) |

*n≠56

Table S6 Baseline values and treatment in predefined subgroups (continued)

|  | **Group 2: Immunosuppression** | | |
| --- | --- | --- | --- |
|  | all (n=16) | Control (n=8) | Plasma (n=8) |
| SARS-CoV-2 Baseline | | | |
| Percent inhibition NeutraLISA (median (p25,p75))* | 7.0 (4.8, 14.1) | 11.2 (5.7, 25.9) | 5.2 (1.4, 7.7) |
| CT value on Day of randomization/Day 1 (Mean +/- SD)* | 23.1 +/-5.2 | 23.2 +/-4.9 | 23.1 +/-5.7 |
| Study assessments | | | |
| Seven point ordinal scale at randomization |  |  |  |
| SPOS =3 | 4 (25.0%) | 1 (12.5%) | 3 (37.5%) |
| SPOS =4 | 9 (56.3%) | 5 (62.5%) | 4 (50.0%) |
| SPOS =5 | 3 (18.8%) | 2 (25.0%) | 1 (12.5%) |
| Laboratory (median (p25,p75)) |  |  |  |
| WBC (Gpt/l) | 8.1 (4.5, 9.8) | 8.9 (4.5, 9.6) | 6.8 (4.7, 10.2) |
| Lymphocytes (Gpt/l)* | 0.5 (0.3, 0.6) | 0.5 (0.2, 0.5) | 0.5 (0.3, 0.8) |
| CRP (mg/l) | 70.7 (43.2, 143.3) | 54.0 (39.6, 103.8) | 117.0 (52.0, 170.7) |
| LDH (U/l) | 419.5 (319.5, 471.5) | 419.5 (319.5, 469.0) | 425.5 (319.0, 511.0) |
| D-Dimer (mg/l)* | 1.2 (0.6, 1.5) | 1.3 (0.6, 1.5) | 0.7 (0.5, 1.2) |
| Troponin (pg/ml)* | 22.8 (10.5, 46.0) | 24.6 (10.5, 111.0) | 17.3 (9.8, 31.0) |
| Treatment (incl. cross-over day 10) | | | |
| Plasma received |  |  |  |
| Convalescent Plasma | 9 | 1 | 8 |
| Convalescent + Vaccinated plasma | 0 | 0 | 0 |
| Vaccinated plasma only | 0 | 0 | 0 |
| Other COVID-19 medication | | | |
| antiinflammatory | 6 (37.5%) | 6 (75%) | 0 (0.0%) |
| small-molecule antiviral | 1 (6.3%) | 1 (12.5%) | 0 (0.0%) |
| biologic antiviral | 0 (0.0%) | 0 (0.0%) | 0 (0.0%) |
| antibiotics | 0 (0.0%) | 0 (0.0%) | 0 (0.0%) |
| anticoagulants | 0 (0.0%) | 0 (0.0%) | 0 (0.0%) |
| other concomitant medication | 1 (6.3%) | 1 (12.5%) | 0 (0.0%) |

*n≠16

Table S6 Baseline values and treatment in predefined subgroups (continued)

|  | **Group 3: Lymphopenia/high D-dimers: aged >50 and ≤75 years** | | |
| --- | --- | --- | --- |
|  | all (n=36) | Control (n=18) | Plasma (n=18) |
| SARS-CoV-2 Baseline | | | |
| Percent inhibition NeutraLISA (median (p25,p75))* | 12.6 (5.6, 36.6) | 14.1 (5.9, 84.5) | 11.0 (5.5, 36.6) |
| CT value on Day of randomization/Day 1 (Mean +/- SD)* | 24.9 +/-4.5 | 24.8 +/-4.1 | 24.9 +/-5.1 |
| Study assessments | | | |
| Seven point ordinal scale at randomization |  |  |  |
| SPOS =3 | 8 (22.2%) | 5 (27.8%) | 3 (16.7%) |
| SPOS =4 | 16 (44.4%) | 8 (44.4%) | 8 (44.4%) |
| SPOS =5 | 12 (33.3%) | 5 (27.8%) | 7 (38.9%) |
| Laboratory (median (p25,p75)) |  |  |  |
| WBC (Gpt/l) | 5.9 (4.3, 7.9) | 6.3 (5.1, 8.6) | 5.2 (3.7, 6.7) |
| Lymphocytes (Gpt/l) | 0.6 (0.4, 0.8) | 0.6 (0.3, 0.8) | 0.7 (0.5, 0.8) |
| CRP (mg/l) | 82.5 (44.1, 133.5) | 97.3 (59.2, 215.7) | 62.3 (35.1, 130.7) |
| LDH (U/l)* | 385.5 (322.0, 491.0) | 439.0 (332.0, 575.0) | 380.0 (295.0, 407.0) |
| D-Dimer (mg/l) | 1.1 (0.6, 1.9) | 1.4 (0.6, 2.2) | 1.1 (0.6, 1.5) |
| Troponin (pg/ml)* | 15.0 (9.1, 28.5) | 13.9 (6.7, 41.6) | 17.0 (13.5, 22.4) |
| Treatment (incl. cross-over day 10) | | | |
| Plasma received |  |  |  |
| Convalescent Plasma | 22 | 4 | 18 |
| Convalescent + Vaccinated plasma | 0 | 0 | 0 |
| Vaccinated plasma only | 0 | 0 | 0 |
| Other COVID-19 medication | | | |
| antiinflammatory | 16 (44.4%) | 7 (38.9%) | 9 (50%) |
| small-molecule antiviral | 5 (13.9%) | 2 (11.1%) | 3 (16.6%) |
| biologic antiviral | 0 (0.0%) | 0 (0.0%) | 0 (0.0%) |
| antibiotics | 4 (11.1%) | 3 (16.6%) | 1 (5.6%) |
| anticoagulants | 2 (5.6%) | 2 (11.1%) | 0 (0.0%) |
| other concomitant medication | 4 (11.1%) | 2 (11.1%) | 2 (11.1%) |

*n≠36

Table S6 Baseline values and treatment in predefined subgroups (continued)

|  | **Group 4: age >75 years** | | |
| --- | --- | --- | --- |
|  | all (n=26) | Control (n=12) | Plasma (n=14) |
| SARS-CoV-2 Baseline | | | |
| Percent inhibition NeutraLISA (median (p25,p75))* | 10.3 (7.1, 26.2) | 7.7 (2.3, 10.8) | 14.3 (9.4, 26.2) |
| CT value on Day of randomization/Day 1 (Mean +/- SD)* | 25.1 +/-5.8 | 24.9 +/-4.3 | 25.3 +/-6.9 |
| Study assessments | | | |
| Seven-point ordinal scale at randomization |  |  |  |
| SPOS =3 | 2 (7.7%) | 1 (8.3%) | 1 ( 7.1%) |
| SPOS =4 | 17 (65.4%) | 7 (58.3%) | 10 (71.4%) |
| SPOS =5 | 7 (26.9%) | 4 (33.3%) | 3 (21.4%) |
| Laboratory (median (p25,p75)) |  |  |  |
| WBC (Gpt/l) | 6.0 (4.6, 8.1) | 6.6 (5.2, 7.8) | 5.6 (4.3, 8.2) |
| Lymphocytes (Gpt/l) | 0.5 (0.4, 0.9) | 0.5 (0.4, 0.9) | 0.6 (0.4, 0.8) |
| CRP (mg/l)* | 72.7 (47.2, 130.3) | 102.0 (47.7, 122.4) | 61.0 (47.2, 137.0) |
| LDH (U/l)* | 350.0 (278.0, 491.0) | 343.0 (275.0, 431.0) | 359.0 (301.0, 524.0) |
| D-Dimer (mg/l)* | 1.1 (0.8, 1.4) | 1.1 (0.6, 1.9) | 1.1 (1.0, 1.3) |
| Troponin (pg/ml)* | 25.3 (15.4, 35.0) | 32.5 (21.6, 56.2) | 17.0 (13.6, 28.7) |
| Treatment (incl. cross-over day 10) | | | |
| Plasma received |  |  |  |
| Convalescent Plasma | 14 | 0 | 14 |
| Convalescent + Vaccinated plasma | 0 | 0 | 0 |
| Vaccinated plasma only | 0 | 0 | 0 |
| Other COVID-19 medication | | | |
| antiinflammatory | 8 (30.8%) | 1 (8.3%) | 7 (50%) |
| small-molecule antiviral | 1 (3.9%) | 0 (0.0%) | 1 (7.1%) |
| biologic antiviral | 0 (0.0%) | 0 (0.0%) | 0 (0.0%) |
| antibiotics | 2 (7.7%) | 0 (0.0%) | 2 (14.3%) |
| anticoagulants | 0 (0.0%) | 0 (0.0%) | 0 (0.0%) |
| other concomitant medication | 2 (7.7%) | 1 (8.3%) | 1 (7.1%) |

*n≠26

Table S7 Adverse Events

|  | Experimental intervention^#^ | Control intervention | Total |
| --- | --- | --- | --- |
| Intensity CTCAE V5* grade |  |  |  |
| - Grade 1 - mild | 118 | 94 | 212 |
| - Grade 2 - moderate | 135 | 151 | 286 |
| - Grade 3 - severe | 51 | 39 | 90 |
| - Grade 4 - life-threatening | 23 | 8 | 31 |
| - Grade 5 - death | 16 | 14 | 30 |
| Maximal Intensity Grade for individual patients |  |  |  |
| - no AEs experienced | 19 ( 24.4%) | 9 ( 16.1%) | 28 ( 20.9%) |
| - Grade 1 - mild | 7 ( 9.0%) | 6 ( 10.7%) | 13 ( 9.7%) |
| - Grade 2 - moderate | 21 ( 26.9%) | 18 ( 32.1%) | 39 ( 29.1%) |
| - Grade 3 - severe | 11 ( 14.1%) | 10 ( 17.9%) | 21 ( 15.7%) |
| - Grade 4 - life-threatening | 5 ( 6.4%) | 4 ( 7.1%) | 9 ( 6.7%) |
| - Grade 5 - death | 15 ( 19.2%) | 9 ( 16.1%) | 24 ( 17.9%) |
| Relation to IMP |  |  |  |
| - Related | 9 | 1 | 10 |
| - Unrelated | 334 | 305 | 639 |

*according to the Common Terminology Criteria for Adverse Events (CTCAE vs5)

#classification according to actual treatment (experimental includes cross-over patients)

AE=adverse events; IMP=Investigational Medicinal Product

| **AEs (MedDRA preferred terms)** | | | | | | | | | |
| --- | --- | --- | --- | --- | --- | --- | --- | --- | --- |
|  | **#AEs under experimental intervention** | **Patients treated with experimental intervention** | **95%-CI** | **#AEs under control intervention** | **Patients treated with control intervention** | **95%-CI** | **#AEs total** | **Total** | **p-value (Chi-squared test)** |
| Patient experiencing at least one AE | 343 | 59 ( 75.6%) | [64.6%; 84.7%] | 306 | 47 ( 83.9%) | [71.7%; 92.4%] | 649 | 106 ( 79.1%) | 0.244 |
| Patient experiencing at least one SAE | 22 | 21 ( 26.9%) | [17.5%; 38.2%] | 19 | 15 ( 26.8%) | [15.8%; 40.3%] | 41 | 36 ( 26.9%) | 0.986 |
| Pyrexia | 29 | 19 ( 24.4%) | [15.3%; 35.4%] | 20 | 14 ( 25.0%) | [14.4%; 38.4%] | 49 | 33 ( 24.6%) | 0.932 |
| Dyspnoea | 17 | 15 ( 19.2%) | [11.2%; 29.7%] | 22 | 18 ( 32.1%) | [20.3%; 46.0%] | 39 | 33 ( 24.6%) | 0.087 |
| Diarrhoea | 8 | 8 ( 10.3%) | [ 4.5%; 19.2%] | 10 | 9 ( 16.1%) | [ 7.6%; 28.3%] | 18 | 17 ( 12.7%) | 0.319 |
| Cough | 7 | 7 ( 9.0%) | [ 3.7%; 17.6%] | 9 | 7 ( 12.5%) | [ 5.2%; 24.1%] | 16 | 14 ( 10.4%) | 0.510 |
| Respiratory failure | 8 | 8 ( 10.3%) | [ 4.5%; 19.2%] | 8 | 4 ( 7.1%) | [ 2.0%; 17.3%] | 16 | 12 ( 9.0%) | 0.534 |
| Anaemia | 6 | 6 ( 7.7%) | [ 2.9%; 16.0%] | 8 | 3 ( 5.4%) | [ 1.1%; 14.9%] | 14 | 9 ( 6.7%) | 0.594 |
| Blood lactate dehydrogenase increased | 9 | 5 ( 6.4%) | [ 2.1%; 14.3%] | 2 | 1 ( 1.8%) | [ 0.0%; 9.6%] | 11 | 6 ( 4.5%) | 0.202 |
| Hypertension | 9 | 9 ( 11.5%) | [ 5.4%; 20.8%] | 2 | 1 ( 1.8%) | [ 0.0%; 9.6%] | 11 | 10 ( 7.5%) | 0.034 |
| Nausea | 5 | 5 ( 6.4%) | [ 2.1%; 14.3%] | 6 | 5 ( 8.9%) | [ 3.0%; 19.6%] | 11 | 10 ( 7.5%) | 0.584 |
| C-reactive protein increased | 5 | 5 ( 6.4%) | [ 2.1%; 14.3%] | 5 | 4 ( 7.1%) | [ 2.0%; 17.3%] | 10 | 9 ( 6.7%) | 0.867 |
| Hypoxia | 4 | 4 ( 5.1%) | [ 1.4%; 12.6%] | 6 | 5 ( 8.9%) | [ 3.0%; 19.6%] | 10 | 9 ( 6.7%) | 0.386 |
| Epistaxis | 3 | 2 ( 2.6%) | [ 0.3%; 9.0%] | 6 | 3 ( 5.4%) | [ 1.1%; 14.9%] | 9 | 5 ( 3.7%) | 0.400 |
| General physical health deterioration | 5 | 5 ( 6.4%) | [ 2.1%; 14.3%] | 4 | 4 ( 7.1%) | [ 2.0%; 17.3%] | 9 | 9 ( 6.7%) | 0.867 |
| Headache | 6 | 3 ( 3.8%) | [ 0.8%; 10.8%] | 3 | 2 ( 3.6%) | [ 0.4%; 12.3%] | 9 | 5 ( 3.7%) | 0.934 |
| Pain | 6 | 5 ( 6.4%) | [ 2.1%; 14.3%] | 2 | 2 ( 3.6%) | [ 0.4%; 12.3%] | 8 | 7 ( 5.2%) | 0.466 |
| Constipation | 2 | 2 ( 2.6%) | [ 0.3%; 9.0%] | 5 | 4 ( 7.1%) | [ 2.0%; 17.3%] | 7 | 6 ( 4.5%) | 0.206 |
| Fatigue | 4 | 4 ( 5.1%) | [ 1.4%; 12.6%] | 2 | 2 ( 3.6%) | [ 0.4%; 12.3%] | 6 | 6 ( 4.5%) | 0.667 |
| Gamma-glutamyltransferase increased | 2 | 2 ( 2.6%) | [ 0.3%; 9.0%] | 5 | 1 ( 1.8%) | [ 0.0%; 9.6%] | 7 | 3 ( 2.2%) | 0.764 |
| Oedema | 5 | 5 ( 6.4%) | [ 2.1%; 14.3%] | 2 | 2 ( 3.6%) | [ 0.4%; 12.3%] | 7 | 7 ( 5.2%) | 0.466 |
| Platelet count decreased | 1 | 1 ( 1.3%) | [ 0.0%; 6.9%] | 6 | 1 ( 1.8%) | [ 0.0%; 9.6%] | 7 | 2 ( 1.5%) | 0.813 |
| Pleural effusion | 3 | 3 ( 3.8%) | [ 0.8%; 10.8%] | 4 | 3 ( 5.4%) | [ 1.1%; 14.9%] | 7 | 6 ( 4.5%) | 0.677 |
| Pneumonia | 3 | 2 ( 2.6%) | [ 0.3%; 9.0%] | 4 | 3 ( 5.4%) | [ 1.1%; 14.9%] | 7 | 5 ( 3.7%) | 0.400 |
| Acute Respiratory Distress Syndrome | 5 | 4 ( 5.1%) |  | 0 | 0 ( 0.0%) |  | 5 | 4 ( 3.0%) | 0.085 |

Table S8 Infusion reactions

| **Infusion reactions** | |
| --- | --- |
|  | **Experimental intervention N=78** |
| Transfusion reaction |  |
| - No | 74 (94.9%) |
| - Transfusion associated circulatory overload (TACO) | 1 (1.3%) |
| - Allergic reaction | 2 (2.6%) |
| - Other | 1 (1.3%) |
| Discontinuation of CP infusion |  |
| - No | 75 (96.2%) |
| - Yes | 3 (3.8%) |

4. **References**

1.	Janssen, M., Schäkel, U., CD Fokou et al., *A Randomized Open label Phase-II Clinical Trial with or without Infusion of Plasma from Subjects after Convalescence of SARS-CoV-2 Infection in High-Risk Patients with Confirmed Severe SARS-CoV-2 Disease (RECOVER): A structured summary of a study protocol for a randomised controlled trial.* Trials, 2020. **21**(1): p. 828.
